# SNORD90 induces glutamatergic signaling following treatment with monoaminergic antidepressants

**DOI:** 10.1101/2023.01.31.23285298

**Authors:** Rixing Lin, Aron Kos, Juan Pablo Lopez, Julien Dine, Laura M. Fiori, Jennie Yang, Yair Ben-Efraim, Zahia Aouabed, Pascal Ibrahim, Haruka Mitsuhashi, Tak Pan Wong, El Cherif Ibrahim, Catherine Belzung, Pierre Blier, Faranak Farzan, Benicio N. Frey, Raymond W. Lam, Roumen Milev, Daniel J. Müller, Sagar V. Parikh, Claudio Soares, Rudolf Uher, Corina Nagy, Naguib Mechawar, Jane A. Foster, Sidney H. Kennedy, Alon Chen, Gustavo Turecki

## Abstract

Most available antidepressants target the serotonergic system, selectively or non-selectively, and yield slow and inconsistent clinical responses, whereas the monoamine changes they elicit do not correlate with treatment response. Recent findings point to the glutamatergic system as a target for rapid acting antidepressants. Investigating different cohorts of depressed individuals treated with serotonergic and other monoaminergic antidepressants, we found that the expression of a small nucleolar RNA, SNORD90, was elevated following treatment response. When we increased SNORD90 levels in the mouse anterior cingulate cortex (ACC), a brain region regulating mood responses, we observed antidepressive-like behaviors. We identified neuregulin 3 (NRG3) as one of the targets of SNORD90, which we show is regulated through the accumulation of N6-methyladenosine modifications leading to YTHDF2 mediated RNA decay. We further demonstrate that a decrease in NRG3 expression resulted in increased glutamatergic release in the mouse ACC. These findings support a molecular link between monoaminergic antidepressant treatment and glutamatergic neurotransmission.

## INTRODUCTION

Antidepressants are the first line treatment for major depressive disorder (MDD), collectively accounting for one of the most prescribed medications (*1*). However, response to antidepressant treatment is variable with less than 50% of patients responding to first trial, and up to 40% with no clinical response after two or more trials (*2*). While most currently available antidepressants act by selectively or non-selectively targeting serotonergic receptors the exact mechanisms by which they effect mood changes and improvements in depression remain unknown. Importantly, although the enhancement of monoamine function, in particular serotonin, can be observed within hours after antidepressant drug administration, clinical improvements are not observed until days or weeks following antidepressant treatment initiation (*3-5*). The delayed clinical response led researchers to study underlying neurobiological adaptations to understand mechanisms of antidepressant response. Monoamines have been a focus of depression studies since most treatment options target this system. However, given the delayed clinical response other neurotransmitter systems have been gaining interest in depression and particularly the glutamatergic system, which is believed to be the target of rapid acting antidepressants (*6, 7*). Moreover, recent evidence suggest that monoaminergic antidepressants may act by modulating the glutamatergic system, although it is still unclear through what molecular mechanisms (*8, 9*).

In this study, we identified a molecular mechanism whereby monoaminergic antidepressants produce an effect on glutamatergic neurotransmission. We found elevated levels of a small nucleolar RNA (snoRNA), SNORD90, in response to antidepressant drug exposure and report that SNORD90 guides N6-methyladenosine (m6A) modifications onto neuregulin 3 (NRG3), which in turn lead to YTHDF2-mediated down-regulation of NRG3 expression and subsequent increases in glutamatergic release.

## RESULTS

### SNORD90 levels are increased in response to monoaminergic antidepressants

We initially examined snoRNA expression using small RNA-sequencing data from peripheral blood samples collected from three independent antidepressant clinical trials (discovery cohort, replication cohort 1 and replication cohort 2) administrating duloxetine, escitalopram, or desvenlafaxine. To identify snoRNAs associated with treatment response, we focused our attention on snoRNAs that were differentially expressed between baseline (T0) and eight weeks after treatment (T8), when clinical outcome was ascertained. In doing so, we identified nine snoRNAs in our placebo controlled double blind discovery cohort that had a significant interaction (p<0.05) between time (T0/T8) and treatment outcome (response/non-response) (figure 1A & supplemental table 1-3). In particular, SNORD90 was the only snoRNA that was consistently up-regulated after antidepressant treatment in subjects who responded to treatment across all three independent cohorts (figure 1B & supplemental figure 1). As such, we further investigated SNORD90.

**Figure 1:**
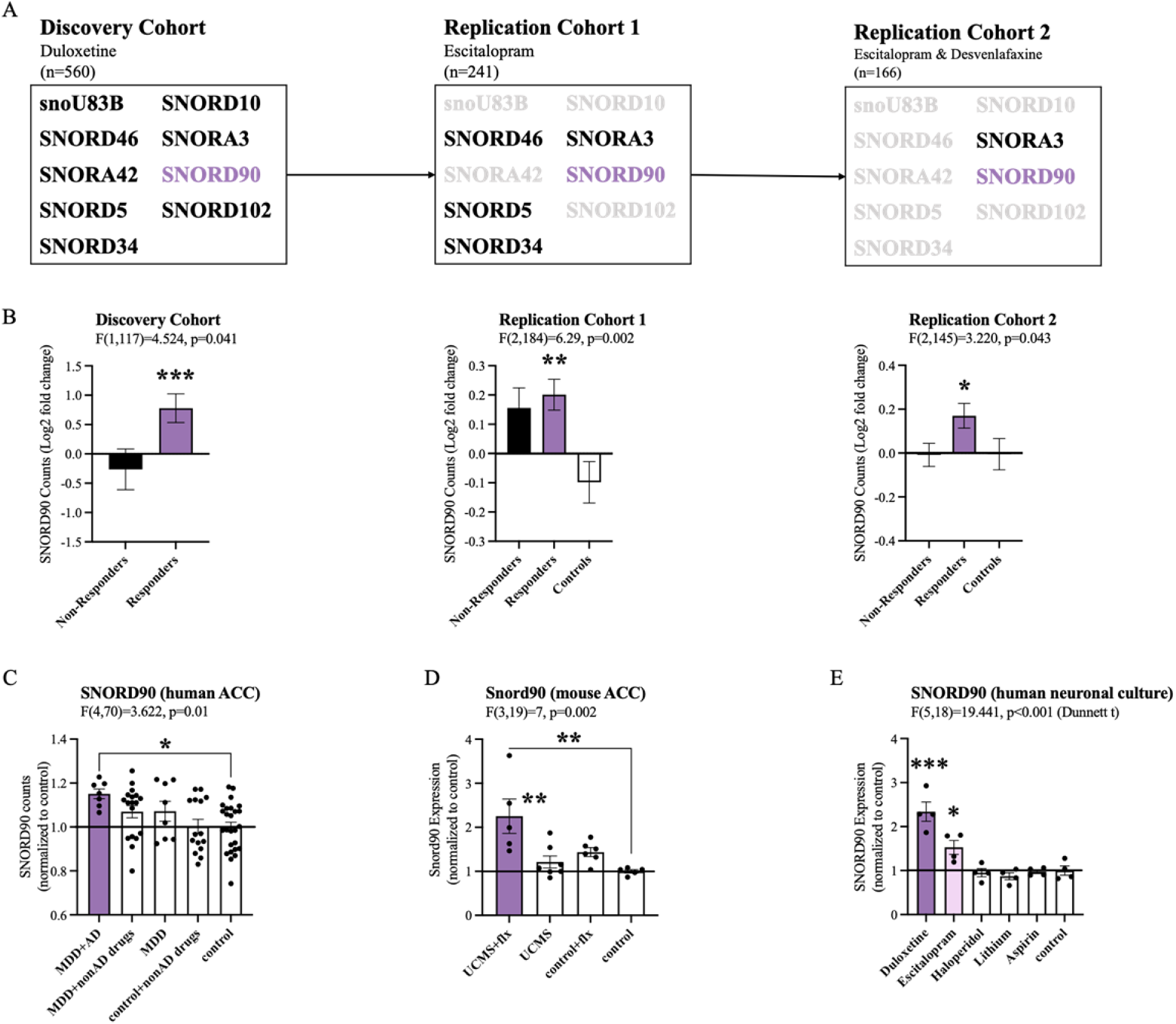
SNORD90 expression is associated with antidepressant treatment response. **A** Two-way mixed multivariable ANOVA indicates a significant interaction between clinical response (responders/non-responders; between-factor) and treatment course (T0/T8; within factor). Nine snoRNAs displayed significant effects in the discovery cohort. Five out of the nine snoRNAs were replicated in replication cohort 1 with SNORD90 and SNORA3 further replicated in replication cohort 2. **B** Log2 fold-change of the expression of SNORD90 before and after antidepressant treatment for all three clinical cohorts. SNORD90 displayed a significantly increased expression after eight weeks of antidepressant treatment specifically in those who responded. **C** Snord90 expression in the ACC of mice that underwent unpredictable chronic mild stress (UCMS) and antidepressant administration. **D** SNORD90 expression in human post-mortem ACC. Samples were separated based on presence or absence of antidepressant drug treatment. **E** SNORD90 expression in human neuronal cultures exposed to various psychotropic drugs. **C-E** Statistical analysis using one-way ANOVA with Bonferroni post-hoc (unless otherwise indicated on the graph). **B-E** All bar plots represent the mean with individual data points as dots. Error bars represent S.E.M. (*p<0.05, **p<0.01, ***p<0.001).

To better understand if SNORD90 expression has similar changes in the brain following antidepressant treatment as observed in peripheral tissue, we investigated human post-mortem anterior cingulate cortex (ACC), a brain region that plays an important role in the regulation of mood (*10, 11*). We studied individuals who died while affected with MDD and were or were not treated with antidepressants. We observed a specific up-regulation of SNORD90 in individuals who were depressed when they died and were actively treated with antidepressants (figure 1C). To follow up these results, we investigated the expression of Snord90 in an unpredicted chronic mild stress (UCMS) mouse model, which is commonly used to study depressive-like behaviors in mice. Mice were subjected to UCMS, followed by administration of fluoxetine (*12*). We specifically profiled the mouse cingulate area 1/2 (cg1/2), a region that is equivalent to the human ACC. Similar to the results observed in humans, we observed a specific up-regulation of Snord90 in the ACC of mice that underwent the UCMS paradigm followed by antidepressant administration, whereas UCMS or antidepressant administration alone did not alter the expression of Snord90 (figure 1D). To assess if the effects observed on the expression of SNORD90 were specific to antidepressants or common to other drugs, we treated human neuronal cells derived from iPSCs and differentiated to a monoaminergic phenotype with several psychotropic drugs, including duloxetine, escitalopram, haloperidol, lithium, or non-psychotropic drugs (aspirin) and regular culture media. We observed that SNORD90 expression was up-regulated exclusively by antidepressant drugs, while other treatments did not significantly alter SNORD90 expression (figure 1E). Together our data suggests that antidepressant treatment response associates with an increase in SNORD90 expression.

### SNORD90 over-expression in mouse ACC induces anti-depressive like behaviours

Given that we observed an upregulation of SNORD90 with antidepressant treatment response in humans and mice, we next investigated if the over-expression of Snord90 in the cg 1/2 cortex of mice has behavioural implications. We over-expressed (OE) Snord90 or a full scrambled Snord90 sequence control in the mouse cg1/2 via bilateral injections of an adeno-associated virus (AAV) followed by a battery of behavioural tests designed to measure anxiety and depressive-like behaviors in mice (figure 2A). We did not observe any differences in the total distance traveled in the open field test, which indicated no general locomotor differences between Snord90 OE and full scramble OE (figure 2B). Snord90 OE increased the amount of time spent in the open arm and decreased the amount of time spent in the closed arm of the elevated plus maze (figure 2C). Moreover, Snord90 OE increased time spent grooming after splashing with 10% sucrose solution and increased time spent struggling in the tail suspension test (figure 2D-E). Lastly, we calculated an integrated emotionality Z-score by combining data from the elevated plus maze, splash test, and tail suspension test (figure 2F). Overall, our results consistently showed that over-expressing SNORD90 levels in cg1/2 yielded decreased emotionality indicative of a decrease in anxiety-like and depressive-like behaviours (figure 2F).

**Figure 2:**
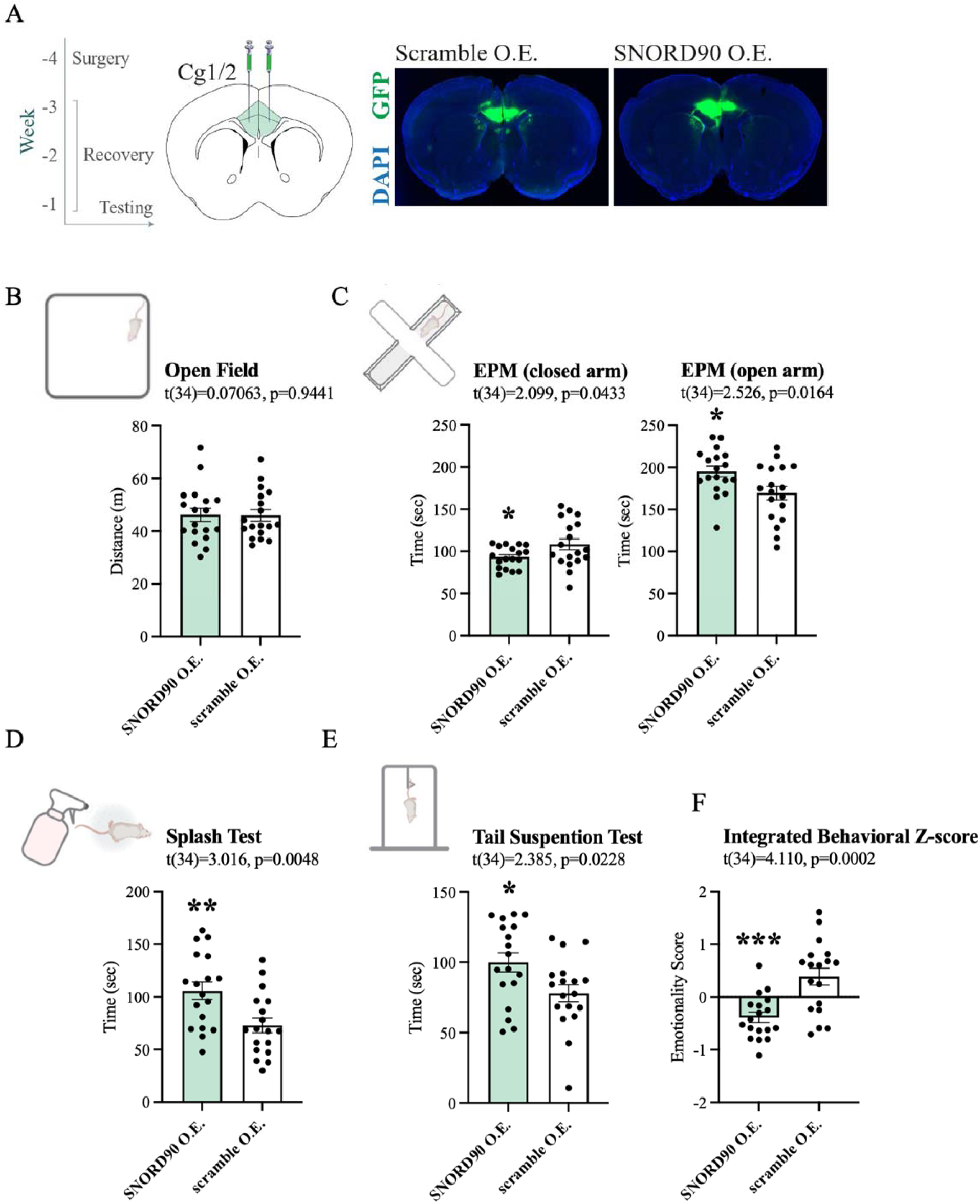
SNORD90 over-expression in mouse Cg1/2 induces anxiolytic and anti depressive-like behaviors **A** Timeline of experimental procedure with time (weeks) in relation to behavioral testing. Surgery for viral injection was performed followed by three weeks of recovery beforebehavioral testing (left). Coronal diagram of the mouse brain representing viral injection site (center). Representative images of GFP expression (green) indicating site specific expression of each construct (right). **B** The open field test showing total distance traveled in meters. **C** The elevated plus maze test with total time spent in the closed and open arms of the maze. **D** The splash test with total grooming time. **E** The tail suspension test with total struggling time. **F** Emotionality z-score integrating the EPM, SPL, and TST. **C-G** Statistical analysis using student’s two-tailed t-test. All bar plots represent the mean with individual data points as dots. Error bars represent S.E.M. (*p<0.05, **p<0.01, ***p<0.001).

### SNORD90 directly down-regulates NRG3

SNORD90 is subcategorized as an orphan snoRNA as it does not have any canonical rRNA, snRNA, or tRNA targets (*13*). However, more recent studies have indicated that some snoRNAs exhibit atypical functioning such as regulation of alternative splicing and modulating expression of mRNA (*14-16*). Thus, we explored possible RNA targets for SNORD90 using the basic local alignment (BLAST) search tool for base complementarity, and using the C/D box snoRNA target prediction tool “PLEXY” (*17*). Using these in-silico methods we identified NRG3 as a putative gene target for SNORD90 (figure 3A & supplementary table 4-5). More specifically, we identified three predicted binding regions for SNORD90 on the NRG3 pre-mRNA (pre-NRG3) (figure 3A & supplementary table 4-5). NRG3 is a growth factor that is part of the neuregulin family and that is highly enriched in the brain and has been previously associated with psychiatric phenotypes (*18-20*). To determine if SNORD90 regulates NRG3, we first assessed NRG3 expression in the same human and mouse ACC samples, as well as in the neuronal cultures described above (supplementary figure 2A-C). We observed a negative correlation between SNORD90 and NRG3 in all three experimental contexts (supplementary figure 2D-F), and interestingly, we observed the most significant differences in direction of fold-change between SNORD90 and NRG3 expression in groups with antidepressant drug exposure (supplementary figure 2G-I).

**Figure 3:**
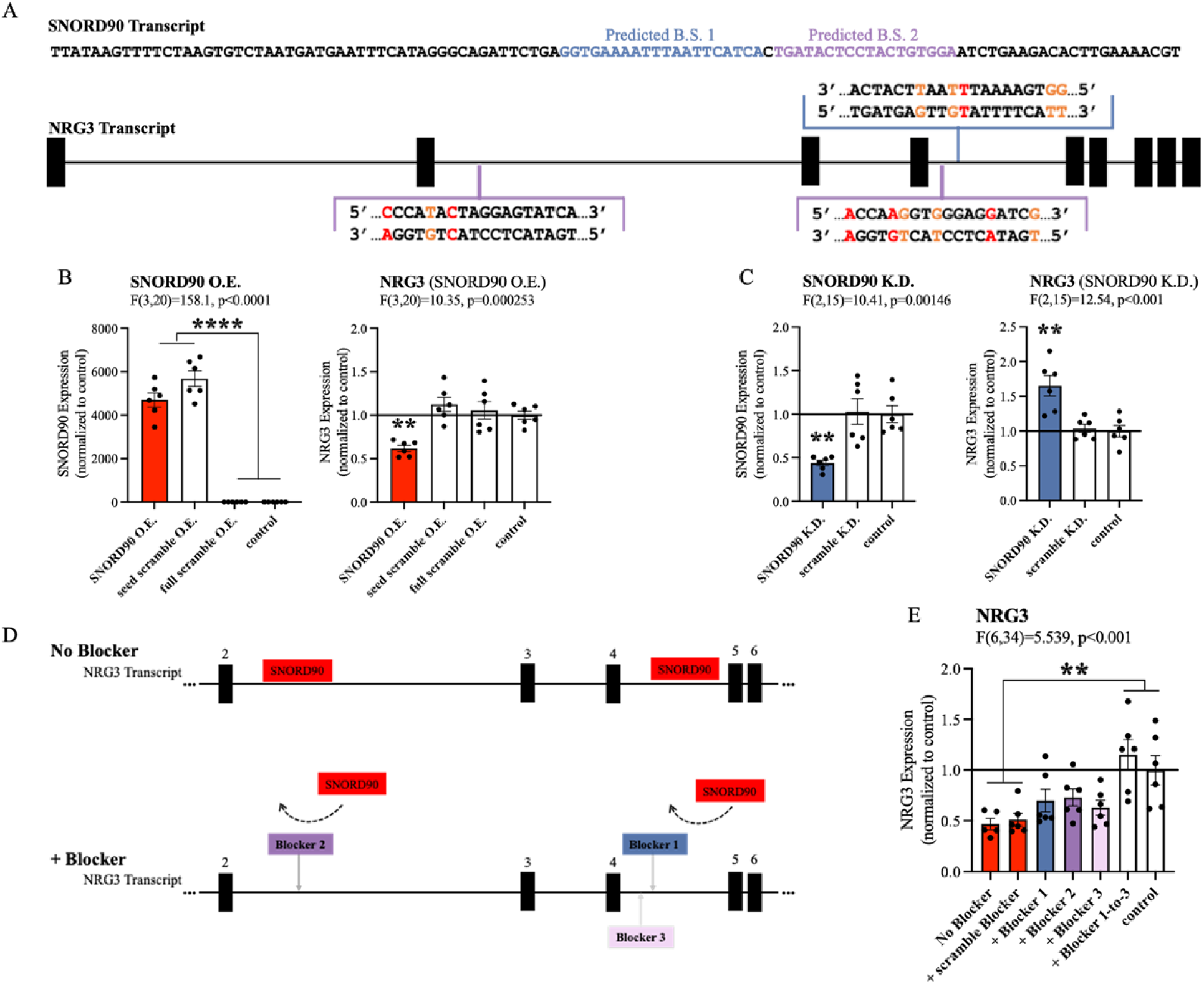
SNORD90 down-regulates NRG3 **A** Full sequence of mature SNORD90 transcript with highlighted regions, labeled predicted B.S. 1 and predicted B.S. 2, which are predicted to bind to NRG3. Schematic representation of NRG3 pre-mRNA transcript indicating regions on NRG3 where SNORD90 is predicted to bind. The color of the bracket corresponds to predicted B.S.-1 or predicted B.S.-2. **B** Expression of SNORD90 (left) and NRG3 (right) after over-expressing SNORD90 or scrambled controls (right). **C** Expression of SNORD90 (left) and NRG3 (right) after knocking-down SNORD90 with antisense oligonucleotides (ASO) and scrambled ASO. **D** Schematic representation of co-transfection of SNORD90 over-expression vector without target blockers (top) and with target blockers (bottom). Target blockers were designed to bind to regions SNORD90 is predicted to bind, consequently blocking SNORD90 from interacting with those regions on NRG3 (bottom). **E** Expression of NRG3 after over-expression of SNORD90 and target blockers. Target blockers were included one site at a time and all three sites together. **B-D, F** All bar plots represent the mean with individual data points as dots. Error bars represent S.E.M. All statistical analysis utilized a one-way ANOVA with Bonferroni post-hoc (*p<0.05, **p<0.01, ***p<0.001).

To further investigate the relationship between SNORD90 and NRG3, we over-expressed SNORD90 in human neural progenitor cells (NPCs) and assessed the resulting effects on NRG3 expression (supplementary figure 3A-B). We constructed AAV vectors expressing wild-type SNORD90 (SNORD90 OE), as well as two scrambled controls (supplementary figure 3A). The first control had a scramble sequence in the central region of the SNORD90 transcript, where the predicted complementary sequence to NRG3 lies (seed scramble OE) and the second control had a full scramble of the entire SNORD90 transcript (full scramble OE) (supplementary figure 3A). SNORD90 OE resulted in a ∼50% decrease in NRG3 expression at both the mRNA and protein levels (figure 3B and supplementary figure3C). Seed scramble OE and full scramble OE did not alter the expression of NRG3, indicating the region of predicted complementarity between SNORD90 and NRG3 plays an important role in the ability of SNORD90 to down-regulate NRG3 (figure 3B). We also measured the expression of pre-NRG3, since our in-silico predictions indicated that SNORD90 is primarily interacting with intronic regions of pre-NRG3, however, pre-NRG3 expression was not altered by SNORD90 OE or any of the scrambled controls (supplementary figure 3D). We next examined the effects of SNORD90 knock-down (KD) using antisense oligonucleotides (ASO) (supplementary figure 4A-B). We screened four ASOs that target different regions of SNORD90 and selected the ASO achieving the best KD (supplemental figure 4A-B). SNORD90 KD by approximately 55% resulted in a ∼50% upregulation of NRG3 expression (figure 3C). We again did not observe any significant changes at the level of pre-mRNA expression (supplementary figure 4C). Finally, to further confirm the importance of direct interaction between SNORD90 and NRG3, we designed target blockers which have sequence complementarity to the predicted SNORD90 binding sites on pre-NRG3 (figure 3D). Target blockers were co-transfected with our SNORD90 OE vectors (figure 3D).

Interestingly, when each individual site was blocked, we observed a partial rescue of NRG3 expression (figure 3E). However, simultaneously blocking all three predicted sites was required for a complete rescue of the downregulation effects of SNORD90 OE (figure 3E). Pre-NRG3 expression was not altered by any target blockers (supplementary figure 5). Together our data suggests that SNORD90 directly down-regulates NRG3 expression through multiple interaction sites on intronic regions.

### SNORD90 associates with RBM15B and guides m6A modifications onto NRG3

We next asked what mechanisms may explain SNORD90’s effects on NRG3 given that it interacts with intronic regions of NRG3, and yet, it does not affect pre-mRNA levels of NRG3. Since SNORD90 displays atypical functioning, we posited that it could regulate NRG3 through the recruitment of a unique set of RNA binding proteins (RBPs) that is different from canonically functioning C/D box snoRNAs (*21*). Using the in-silico tool “oRNAment” we identified a sequence-motif for RNA Binding Motif Protein 15B (RBM15B); this motif sequence was further confirmed by Van Nostrand et al., 2020 (*22, 23*) (supplemental table 6). RBM15B is a key regulator of RNA N6-methyladenosine (m6A) modifications by facilitating interactions with Wilms’ tumor 1-associating protein (WTAP), which in turn binds to methyltransferase like 3 (METTL3) forming a major m6A writer complex (*24*). To confirm the association of SNORD90 with RBM15B, we performed RNA immunoprecipitation (RIP) against RBM15B as well as the canonical core enzymatic protein for C/D snoRNAs, Fibrillarin (figure 4A) (*25, 26*). We observed a significantly higher level of association of SNORD90 to RBM15B compared to fibrillarin whereas a canonically functioning snoRNA, SNORD44, displayed enrichment for fibrillarin IP (figure 4A). Furthermore, when we over-expressed SNORD90 we observed an increase in association of SNORD90 with RBM15B, compared to over-expression of a scrambled control (figure 4B). We did not observe an increase of SNORD90 association with Fibrillarin following SNORD90 OE, indicating that SNORD90 showed preference for interacting with RBM15B (figure 4B). Since snoRNAs and RBM15B are both found in the nucleus this may in part explain SNORD90’s role in interacting with intronic regions of NRG3 pre-mRNA (*21, 24*). Canonical snoRNAs function by guiding their partner proteins to target transcripts which, in-turn, induce a chemical modification (*27*). Thus, we hypothesized that SNORD90 is likely acting as a guide RNA for RBM15B and its associated m6A writer complex, resulting in an increase in m6A levels on NRG3. To test this hypothesis, we measured total m6A abundance on NRG3 and pre-NRG3 transcripts from our SNORD90 OE in-vitro NPC culture experiments detailed above. We observed an increase of m6A abundance on both NRG3 and pre-NRG3, following SNORD90 OE, whereas seed scramble OE and full scramble OE did not alter m6A abundance (figure 4C-D). Furthermore, introducing target blockers blunted the increase of m6A abundance on both NRG3 and pre-NRG3 (figure 4E-F). To further test RBM15B’s role in the observed increase in m6A levels on NRG3, we used dicer-substrate short interfering RNAs (dsiRNAs) to knock-down RBM15B (RBM15B KD) (supplementary figure 6). RBM15B KD followed by SNORD90 OE blunted the increase of m6A levels on NRG3 and pre-NRG3 (figure 4G-H). RBM15B KD also blunted the decrease of NRG3 expression following SNORD90 OE (figure 4G-H). Interestingly we observed a significant negative correlation between m6A levels and expression of NRG3, but this was not observed for pre-NRG3, which could possibly be explained by the fact that m6A-readers are primarily located in the cytoplasm, and thus would only recognize the m6A-modifications of the mature NRG3 transcript and promote its decay when it is shuttled out of the nucleus into the cytoplasm (supplementary figure 7). Ke et al., 2017 demonstrated that m6A modifications are added onto nascent pre-mRNA during transcription (*28*). Moreover, m6A levels remain unchanged between nascent pre-mRNA and steady-state mRNA in the cytoplasm (*28*). Our results further support these findings as it is possible that SNORD90 is guiding m6A modifications onto NRG3 at the pre-mRNA level, which are retained in the mRNA molecule. Although it is unclear why SNORD90 targets intronic regions, while it seems that m6A modifications are deposited onto exonic regions, this could perhaps be explained by the secondary structure of the RNA molecule where exonic regions could be looping closer to the SNORD90-guided methylation complex.

**Figure 4:**
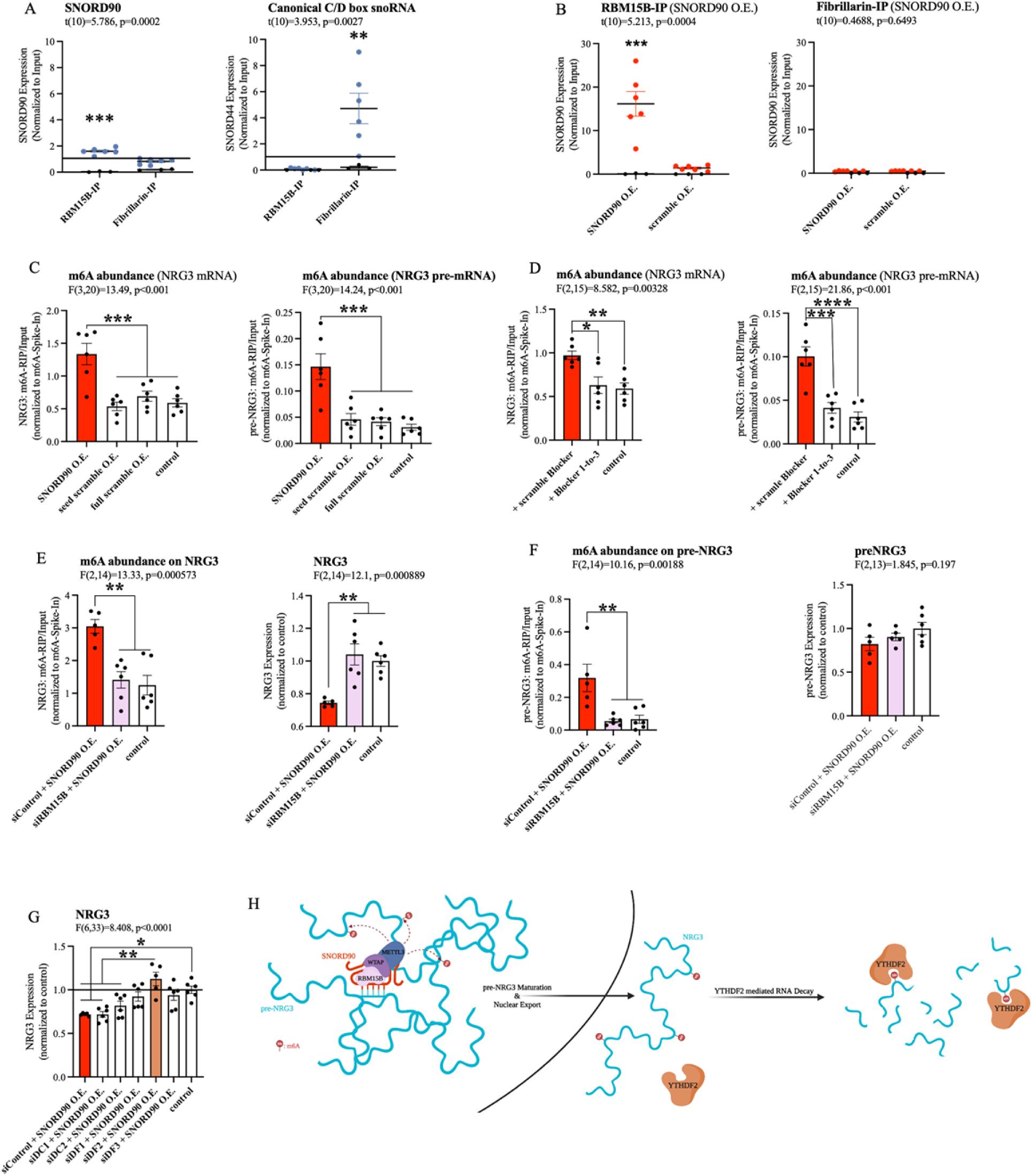
SNORD90 is a guide RNA for RBM15B and increases m6A abundance on NRG3 **A** Abundance of SNORD90 (left) and canonical snoRNA (SNORD44; right) in RBM15B-IP and fibrillarin-IP fractions. **B** Abundance of SNORD90 in RBM15B-IP (left) and fibrillarin-IP (right) following over-expression of SNORD90 or scramble control. **C** Abundance of m6A modifications on NRG3 mRNA (left) and NRG3 pre-mRNA (right) following over-expression of SNORD90 or scramble controls. **D** Abundance of m6A modifications on NRG3 mRNA (left) and NRG3 pre-mRNA (right) following SNORD90 overexpression with target blockers. **E** Abundance of m6A modifications on NRG3 (left) and NRG3 expression (right) following RBM15B knock-down (siRBM15B) or negative control (siControl) and SNORD90 over-expression. **F** Same as E but on NRG3 pre-mRNA. **G** Expression of NRG3 after knocking-down m6A reader proteins followed by over-expression of SNORD90. **H** Schematic overview of SNORD90 regulation of NRG3 expression. SNORD90 interacts with m6A writer complex in the nucleus and guides this complex onto NRG3 increasing m6A abundance. The increase in m6A abundance is not recognized until NRG3 reaches the cytoplasm where it undergoes YTHDF2 mediated RNA decay. All bar plots represent the mean with individual data points as dots. Error bars represent S.E.M. Statistical analysis utilized as follows: **A-B** Student’s two-tailed t test. **E** Pearson correlation. **C-D, F-H** One-way ANOVA with Bonferroni post-hoc (*p<0.05, **p<0.01, ***p<0.001).

The YTH family of proteins are the best described readers of RNA m6A modifications. The YTH family includes: YTHDF1, YTHDF2, YTHDF3, YTHDC1, and YTHDC2. Of these, YTHDF1, YTHDF2 and YTHDF3 regulate RNA stability, whereas functional implications for YTHDC1 and YTHDC2 are less well defined (*29, 30*). Furthermore, YTHDF1, YTHDF2, YTHDF3, and YTHDC2 are primarily located in the cytoplasm, whereas YTHDC1 is primarily located in the nucleus (*31*). We selectively knocked-down each of the above-mentioned m6A-readers using dsiRNAs followed by SNORD90 OE to elucidate which m6a-reader(s) are involved in the down-regulation of NRG3 (figure 4I & supplementary figure 8). Although YTHDF1 KD, YTHDF2 KD, and YTHDF3 KD all showed the ability to blunt the decreased NRG3 expression following SNORD90 OE, YTHDF2 displayed the most robust ability to blunt this effect, recovering NRG3 expression to WT patterns (figure 4I). On the other hand, YTHDC2 KD and YTHDC1 KD did not blunt the decrease of NRG3 expression after SNORD90 OE; displaying similar patterns of NRG3 expression to SNORD90 OE with a dsiRNA scramble control (siControl) (figure 4I). Interestingly, YTHDC1 KD, the nuclear m6a-reader, resulted in a decrease of pre-mRNA levels of NRG3 expression compared to WT, while knock-down of all other m6a-readers had no significant effect on pre-NRG3 expression levels (supplementary figure 9). Together this evidence supports the hypothesis that 1) SNORD90 increases NRG3 m6A transcript methylation and subsequent decay, and 2) that the decay occurs primarily in the cytoplasm (figure 4J). This, in turn, explains why we only observed changes in NRG3 mRNA and protein levels, but did not observe changes in pre-mRNA levels following SNORD90 OE.

### SNORD90 mediates down-regulation of Nrg3 resulting in increased glutamatergic neurotransmission

We next investigated the functional relevance of decreased NRG3 expression in-vivo. NRG3 has previously been shown to interact with syntaxin, disrupting SNARE complex formation in the presynaptic terminal, and inhibiting vesicle docking (*19*). Nrg3 knock-out resulted in increased probability of glutamate release in mouse hippocampal neurons (*19*). To investigate if SNORD90-mediated down-regulation of NRG3 has a similar effect in mice, we first investigated if SNORD90’s ability to down-regulate NRG3 is conserved in mice. We performed in-silico target prediction between mouse Snord90 and Nrg3 using “PLEXY” (supplementary table 7).

Although predicted sequence complementary sites between Snord90 and Nrg3 are not as robust as in humans, there is conservation between the two species (supplementary table 7 and supplementary figure 10). To further explore this effect, we over-expressed Snord90 or a full scrambled control in the mouse ACC (cg1/2) via bilateral injections of an AAV virus (figure 5A). RT-qPCR confirmed successful over-expression of Snord90 and subsequent down-regulation of Nrg3 in mice (figure 5B). Next, we performed whole-cell patch-clamp recordings from pyramidal neurons of the ACC in acute brain slices from mice over-expressing Snord90 or the full scramble control. We observed an increase in spontaneous excitatory postsynaptic currents (sEPSC) frequency following Snord90 OE compared to full scramble control without any effect on sEPSC amplitude (figure 5C-E). This increase in glutamatergic neurotransmission is likely due to an increase in glutamate release probably resulting from NRG3 pre-synaptic expression and effect on SNARE complex (*19*). Together this indicates that SNORD90-mediated down-regulation of NRG3 has implications in glutamate neurotransmission, which translates to behavioral changes such as anxiolytic and anti-depressive-like behaviors.

**Figure 5:**
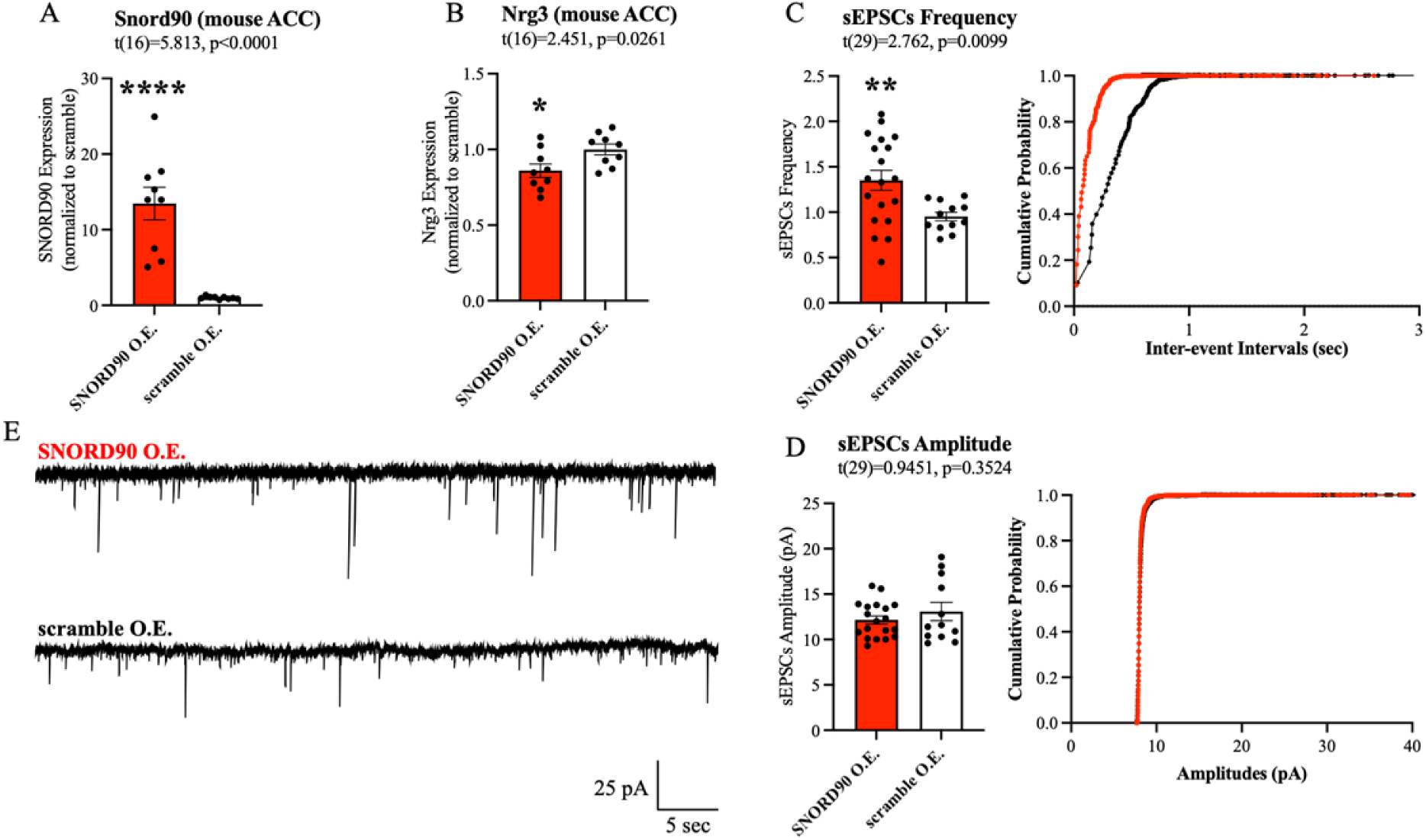
SNORD90 induced down-regulation of Nrg3 increases glutamatergic neurotransmission **A** qPCR confirmation of Snord90 over-expression in cg1/2. **B** Nrg3 expression after Snord90 over-expression. **C-E** Whole-cell patch-clamp recordings in Cg1/2 acute brain slices from mice over-expressing SNORD90 or scramble control. sEPSCs were recorded from pyramidal neurons at -70 mV. **A-C** & **E** Statistical analysis using student’s two-tailed T-test. All bar plots represent the mean with individual data points as dots. Error bars represent S.E.M. (*p<0.05, **p<0.01, ***p<0.001).

## DISCUSSION

In this study, we demonstrated that SNORD90 mediates antidepressant drug action through regulation of NRG3. While it is unclear how serotonergic antidepressant drugs may promote the increase of SNORD90 expression, one possible mechanism may be through histone serotonylation. Previous studies have demonstrated that serotonin can covalently attach to histone H3 and influence gene expression (*32*). The administration of serotonergic antidepressants could change intracellular monoamine levels influencing the expression of SNORD90; however, future studies are needed to investigate this hypothesis. Our results indicate that SNORD90 associates with RBM15B and is involved in mediating m6A modifications, specifically onto NRG3. This is a slight but significant divergence from canonically functioning C/D box snoRNAs that are known to associate with fibrillarin, a methyltransferase responsible for 2’O-methylation (2’OMe) (*33, 34*). Whereas 2’OMe can occur on the ribose of any base and is associated with increased RNA stability, m6A is an adenosine specific modification with a diverse range of effects on RNA stability (*31, 35*). Specifically, our data show that increased m6A levels elicit NRG3 decay through recognition by the m6A reader YTHDF2. Furthermore, SNORD90’s influence on NRG3 appears to bridge a relationship between the monoaminergic and the glutamatergic systems. Many studies have shown that antidepressant drugs target the monoaminergic system but also affect the glutamatergic system (*9*). However, it has not been clear until now what is mediating these effects, though they have often been attributed to off target binding of antidepressant drugs. Our study offers molecular evidence supporting one particular mechanism that might mediate the link between serotonin targeting drugs and activation of the glutamatergic system. Although NRG3 expression has been associated with psychiatric disorders including MDD, it has not previously been associated with antidepressant treatment (*18*). NRG3 is primarily expressed in excitatory pyramidal neurons, and accordingly, the increase of SNORD90 in response to antidepressant treatment, can yield specific effects on excitatory glutamatergic neurons and thus glutamatergic neurotransmission (*19, 36*). Gaining a better understanding of the molecular mechanisms of antidepressant treatment response will offer new targets for the development of more effective treatments of MDD.

## MATERIALS AND METHODS

### Human Clinical trial Subjects

Three independent cohorts were used in this study, comprising 660 individuals (*37-39*).

Cohort 1 (N=258) was obtained in collaboration with Lundbeck A/S sponsored clinical trials and is composed of individuals diagnosed with MDD in a current major depressive episode (MDE) who were enrolled in a double-blind clinical trial and received treatment with either duloxetine (60mg), a serotonin-norepinephrine reuptake inhibitor (SNRI), or placebo for eight weeks. For each patient, peripheral blood samples were collected at baseline (T0) and after treatment (T8). Participants, aged 19–74 years, were recruited based on a primary diagnosis of MDD and MDE lasting at least 3 months, with a severity score on the Montgomery-Åsberg Depression Rating Scale (MADRS) of ≥22 at T0. Participants resistant to at least two previous AD treatments or who had received electroconvulsive therapy in the 6 weeks before the study began were excluded. Other exclusion criteria included: MDE in bipolar disorder, presence of psychotic features, and recent substance use disorder. This clinical trial was approved by ethics boards of participating centers, and all participants provided written informed consent. www.ClinicalTrials.gov (11984A NCT00635219; 11918A NCT00599911; 13267A NCT01140906). For more details, please refer methods section of Lopez et al., 2017 (*37*).

Cohort 2 (N=236) was obtained in collaboration with the Canadian Biomarker Integration Network in Depression (CAN-BIND) and is composed of individuals diagnosed with MDD (N=153) and healthy controls (N=83). Patients and controls were recruited at six Canadian clinical centers. Exclusion criteria included personal and family history of schizophrenia or bipolar disorder, or current substance dependence. Depressed patients were treated with escitalopram (10-20mg per day), a selective serotonin reuptake inhibitor (SSRI), for eight weeks.

Depression severity was assessed at baseline and after treatment by MADRS. This trial was approved by ethics boards of participating centers and all participants provided written informed consent. www.ClinicalTrials.gov identifier NCT01655706. Registered 27 July 2012. For more details, please refer to Kennedy et al., 2019 (*38*).

Cohort 3 (N=166) was obtained at the Douglas Mental Health University Institute and is composed of healthy controls (N=28) and individuals diagnosed with MDD (N=138) who were enrolled in the community outpatient clinic at the Douglas Mental Health University Institute. Depressed patients were treated with either desvenlafaxine, a serotonin-norepinephrine reuptake inhibitor (SNRI) or escitalopram, a selective serotonin reuptake inhibitor (SSRI). For each patient, a blood sample was taken at baseline before treatment administration and 8 weeks post-treatment. Participants (healthy controls and individuals with MDD) were excluded from the study if they had comorbidity with other major psychiatric disorders, if they had positive tests for illicit drugs at any point during the study, or if they had general medical illnesses. Individuals with MDD were not receiving antidepressant treatment at the onset of the trial, and received a diagnosis of MDD without psychotic features, according to the Statistical Manual of Mental Disorders, Fourth Edition (DSM-IV). Control subjects were excluded if they had a history of antidepressant treatment. Eligible participants were randomized to either desvenlafaxine (50-100mg) or escitalopram (10-20mg) treatment. All subjects included in the study provided informed consent, and the project was approved by The Institutional Review Board of the Douglas Mental Health University Institute. For more details, please refer to Jollant et al., 2020 (*39*).

#### Clinical Assessment

All participants from all cohorts were assessed for depression severity after 6 to 8 weeks of treatment. To quantify treatment response, we calculated percentage change of MADRS scores (from baseline to after treatment). We used percentage change to correct for the potential effects of differential baseline scores. Additionally, we classified participants as responder/non-responder based on >50% decrease in MADRS scores from baseline.

#### Human peripheral blood sample processing and RNA Extraction

Peripheral blood samples from cohort 1 were collected in PAXgene blood RNA tubes (PreAnalytix). Total RNA was isolated from whole blood using the PAXgene Blood miRNA Kit (Qiagen, Canada) according to the manufacturer’s instructions. Peripheral blood samples from cohort 2 and cohort 3 were collected in EDTA blood collection tubes and passed through LeukoLOCK filters (ThermoFisher) to capture the total leukocyte population, eliminating red blood cells, platelets, and plasma. Filters, containing leukocytes, were frozen at -80C for storage until ready for sample processing. Total RNA was extracted using a modified version of the LeukoLOCK Total RNA Isolation System protocol (ThermoFisher). All samples were treated with DNase digestion during RNA purification using the RNase-Free DNase kit (Qiagen). RNA yield and quality were determined using the Nanodrop 1000 (Thermo Scientific, USA) and Agilent 2200 Tapestation (Agilent Technologies, USA).

#### Library Construction and Small RNA-Sequencing

Libraries for cohort 1 and cohort 3 were prepared using the Illumina TruSeq Small RNA protocol following the manufacturer’s instructions. Libraries for cohort 2 were prepared using NEB small RNA protocol following manufacturer’s instructions. All libraries were purified using biotinylated magnetic AMPure beads that allow for selection of specified complementary cDNA products bound to streptavidin. A total of 50μl of amplified cDNA were mixed and purified twice with AMPure XP beads in a 1.8:1 ratio (beads/sample). Cohort 1 was sequenced using Illumina HiSeq2500, cohort 2 was sequenced using Illumina HiSeq4000, and cohort 3 was sequenced using Illumina HiSeq2000. All samples were sequenced at the McGill University and Genome Quebec Innovation Centre (Montreal, Canada) using 50 nucleotide single-end reads. All sequencing data was extracted from FASTQ files and processed using CASAVA 1.8+. Illumina adapter sequences were trimmed using the Fastx_toolkit, and additionally filtered by applying the following cut-offs: (1) Phred quality (Q) score higher than 30, (2) reads between 15-40nt in length, (3) adapter detection based on perfect-10nt match, and (4) removing reads without detected adapters. Bowtie35 (John Hopkins University) was used to align reads to the human genome (GRCh37). Furthermore, Rfam database was used to map reads to known small nucleolar RNAs. Sequencing data was normalized with the Bioconductor-DESeq2 package.

#### Statistical analysis

For each subject in these trials, samples collected before administration of an antidepressant (T0) and eight weeks following antidepressant treatment (T8) were analyzed according to response to treatment based on MADRS score changes. For the discovery cohort, antidepressant treated, and placebo treated subjects were analyzed separately.

A two-way mixed multivariable analysis of variance (2WM-MANOVA) was used to identify snoRNAs that had a significant interaction between treatment response (response/non-response) and treatment course (T0/T8). For cohort 1, antidepressant treated, and placebo treated subjects were analyzed separately. All detected snoRNAs were assessed in cohort 1. Only snoRNAs that showed significant interactions were assessed in cohort 2 and subsequently only snoRNAs that were replicated in cohort 2 were assessed in cohort 3. For all cohorts, outliers were identified using boxplot methods with values above quartile 3 (Q3) + 1.5 interquartile range (IQR) or below quartile 1(Q1) - 1.5IQR (IQR=Q3-Q1). Shapiro-Wilk test and QQ plots were used to assess data normality. All above mentioned tools were apart of publicly available R package “rstatix” (https://CRAN.R-project.org/package=rstatix) (*40*).

##### Unpredicted chronic mild stress (UCMS) mouse model

Eight-week old male BALB/c mice (N=23; Centre d’Elevage Janvier, Le Genest St. Isle, France) were divided into four groups as described by Herve et al., 2017 (*12*). In brief, control group (N = 5) was kept in standard housing conditions for 8 weeks. UCMS only group (N = 7) comprised of mice subjected to the Unpredictable Chronic Mild Stress (UCMS) procedure for 8 weeks. UCMS-flx group (N=5) included mice that were subjected to the UCMS procedure for 8 weeks and treated in parallel by fluoxetine (flx) during the last 6 weeks. Flx only group (N = 6) included mice that did not undergo the UCMS procedure but were treated with fluoxetine during the last 6 weeks. Mice from the control and flx only groups were housed in standard cages, whereas the UCMS-exposed mice were isolated in individual home cages with no physical contact with other mice. The stressors used were varied and applied in a different sequence each week to avoid habituation.

Stressors consisted of housing on damp sawdust (about 200 mL of water for 100 g of sawdust), sawdust changing (replacement of the soiled sawdust by an equivalent volume of new sawdust), placement in an empty cage (usually the home cage of the subject, but with no sawdust), placement in an empty cage with water (the mouse is placed in its empty cage, whose bottom has been filled up with 1 cm high water at 21°C), switching cages (also sometimes termed as social stress: the mouse from a cage A is placed in the soiled cage from mouse B, mouse B itself being absent in order to avoid aggressive interactions), cage tilting (45°), predator sounds, introduction of rat or cats feces as well as fur in the mouse home cage, inversion of the light/dark cycle, lights on for a short time during the dark phase or light off during the light phase, confinement in small tubes (diameter: 4 cm; length: 5 cm).

#### Mice Behavior

Weight and coat state were measured weekly, as markers of UCMS-induced depressive-like behavior, except for the last week before sacrifice, when coat state from seven different areas of the body was recorded twice, separated by 3-day intervals. At the end of the 8th week, a complementary test of nest building was performed just before sacrifice. The test was administered by isolating mice in their home cages. For additional details please refer to Herve et al., 2017 (*12*).

#### Quantification and statistical analysis

Details related brain dissection and RNA extraction can be found at Herve et al., 2017 (*12*). RNA was reverse-transcribed using M-MLV Reverse Transcriptase (200 U/µL) (ThermoFisher) with random hexamers. SNORD90 and NRG3 were quantified by RT-PCR using SYBR green (Applied Biosystems). Reactions were run in triplicate using the QuantStudio 6 Flex System and data collected using QuantStudio Real-Time PCR Software v1.3. One-way ANOVA was used as described above.

##### Human Post-Mortem Brain

Post-mortem samples of dorsal ACC (Brodmann Area 24) were obtained, in collaboration with the Quebec Coroner’s Office, from the Douglas-Bell Canada Brain Bank (Douglas Mental Health University Institute, Montreal, Quebec, Canada). Groups were matched for post-mortem interval (PMI), pH and age. Psychological autopsies were performed as described previously, based on DSM-IV criteria (*41*). The control group had no history of major psychiatric disorders. All cases met criteria for MDD or depressive disorder not-otherwise-specified. Written informed consent was obtained from next-of-kin. This study was approved by the Douglas Hospital Research Centre institutional review board.

#### RNA-sequencing

RNA was extracted from all brain samples using a combination of the miRNeasy Mini kit and the RNeasy MinElute Cleanup kit (Qiagen), with DNase treatment, and divided into small (<200 nt) and large (>200 nt) fractions. RNA quality, represented as RNA Integrity Number, was assessed using the Agilent 2200 Tapestation. Small RNA-seq libraries were prepared from the small RNA fraction, using the Illumina TruSeq Small RNA protocol following the manufacturer’s instructions. Samples were sequenced at the McGill University and Genome Quebec Innovation Centre (Montreal, Canada) using the Illumina HiSeq2000 with 50nt single-end reads. All sequencing data were processed using CASAVA 1.8 + (Illumina) and extracted from FASTQ files. The Fastx_toolkit was used to trim the Illumina adapter sequences. Additional filtering based on defined cutoffs was applied, including: (1) Phred quality (Q) mean scores higher than 30, (2) reads between 15 and 40 nt in length, (3) adapter detection based on perfect-10nt match, and (4) removal of reads without detected adapter. In addition, we used Bowtie [24] to align reads to the human genome (GRCh37). Furthermore, all sequencing data was normalized with the Bioconductor—DESeq2 package, using a detection threshold of 10 counts per snoRNA. We retained all snoRNAs with >10 reads in 70% of either group (controls, cases) for differential analyses. RNA extractions, sequencing, and data processing were conducted by blinded investigators. For additional details please refer to Fiori et al., 2020 (*42*).

#### Statistical analysis

Using toxicology screens, each sample was separated into the following groups: MDD with presence of antidepressants, MDD with presence of non-antidepressant drugs, MDD with negative toxicology screen, controls with non-antidepressant drugs, and controls with negative toxicology screens. No control samples were positive for antidepressant drugs. For this analysis we only investigated the expression for SNORD90 from this small RNA sequencing dataset. NRG3 was measured via qPCR from cDNA converted from the same RNA aliquot for each sample. Analysis was performed in R using “rstatix” as mentioned above employing a one-way ANOVA. Shapiro-Wilk test and QQ plots were used to assess data normality.

##### Human hindbrain NPC Culture and neuronal differentiation

Monoamine producing neurons were generated from human induced pluripotent stem cells (iPSCs), using a protocol adapted from Lu et al. 2016 (*43*). Human iPSCs were first cultured in DMEM/F12 (Gibco) supplemented with N2 (Gibco), B27 (Gibco), nonessential amino acids (Gibco), 1% GlutaMAX (Gibco), 2 uM SB431542 (STEMCELL Tech.), 2uM DMH1 (Tocris), and 3uM CHIR99021 (Tocris); collectively referred to as SDC media. Culturing in SDC media for 1 week induces human iPSC differentiation into rostral hindbrain neural stem cells (NSCs). Rostral hindbrain NSCs colonies were selected and re-plated in SDC media supplemented with 1000 ng/ml of SHH C25II (GenScript). Hindbrain NPCs were GBX2, HOXA2, and HOXA4 positive as assessed via quantitative RT-PCR to confirm hindbrain specificity at this developmental stage. Ventral rostral hindbrain NSC colonies were collected and re-plated in SDC + SHH media along with 10ng/ml of FGF4 (PeproTech). SDC + SHH + FGF4 media will induce ventral rostral hindbrain NSC differentiation into neural progenitor cells (NPCs) after 1 week. NPCs were expanded in SDC+SHH+FGF4 media and differentiated into monoamine producing neuron-like cells. NPCs were differentiated, for 1 month, into neuron-like cells in neurobasal media (Gibco) supplemented with N2, B27, NEAA, 1ug/ml laminin (Sigma), 0.2mM vitamin C (Sigma), 2.5uM DAPT (Sigma), 10ng/ml GDNF (GenScript), 10ng/ml BDNF (GenScript), 10ng/ml insulin-like growth factor-I (Pepro Tech), and 1ng/ml transforming growth factor β3 (Pepro Tech). Post-differentiation, neuron-like cells underwent high-performance liquid chromatography (HPLC) to test for monoamine production. HPLC confirmed production of norepinephrine (NE), epinephrine (epi), dopamine (DA), and serotonin (5-HT). All NPCs and differentiated NPCs were seeded on culture plates coated with 100ug/mL poly-L-ornithine (sigma) and 10ug/mL laminin (Sigma) and grown in a 5% CO_2_ humidified incubator at 37°C.

#### High Performance Liquid Chromatography

Cells were sonicated in 60uL of 0.25 N perchloric acid. Protein and cellular debris were cleared by centrifugation at 11,000g at 4°C for 10 min. Pellets were re-suspended in 100μl of 0.1N NaOH. 20uL of tissue homogenates, cleared of protein and debris, was injected using a refrigerated ultiMate 3000 rapid separation autosampler (ThermoFisher) into a HPLC system consisting of a luna 3 u C18 (2) 100 A 75 X 4.6 mm phenomemex and a coulometric electrochemical detector (ThermoFisher) to quantify monoamines. Oxidation and reduction electrode potentials of the analytical cell (5014B; ThermoFisher) were set to +300 and −250 mV respectively. The mobile phase consisting of 73.4mM sodium acetate trihydrate, 66.6mM of citric acid monohydrate, 0.025mM Na2EDTA, 0.341mM 1-octanesulfonic acid, 0.71mM of Triethylamine and 6% (v/v) methanol (pH adjusted to 4.0-4.1 with acetic acid) was pumped at 1.5 ml/min by a solvent delivery module (dionex ultimte 3000 rs pump).

##### NPC Drug Treatment

Human NPCs were screened for cytotoxic effects using the MTT assay, and antidepressants were applied at nontoxic concentrations as described by Lopez et al., 2014 (*44*). NPCs were cultured in 24 well plates and differentiated, for 2 weeks, into neuron-like cells in neurobasal media (Gibco) supplemented with N2, B27, NEAA, 1 µg/ml laminin (Sigma), 0.2 mM vitamin C (Sigma), 2.5 µM DAPT (Sigma), 10 ng/ml GDNF (GenScript), 10 ng/ml BDNF (GenScript), 10 ng/ml insulin-like growth factor-I (Pepro Tech), and 1 ng/ml transforming growth factor β3 (Pepro Tech). Following 2 weeks of differentiation, culture media was supplemented with escitalopram (Sigma-Aldrich, E4786; 100 µM), duloxetine (Sigma-Aldrich, Y0001453; 10 µM), haloperidol (Sigma-Aldrich, H1512; 10 µM), lithium (Sigma-Aldrich, L4408; 1 mM), Aspirin (Sigma-Aldrich, A5376; 1mM), or left untreated (controls). Cells for each drug treatment were incubated for 48 h before harvest and RNA extractions. Each drug treatment was performed in triplicate. RNA was extracted using the Zymo DirectZol RNA Extraction kit. cDNA construction and RT-qPCR were as described above. One-way ANOVA analysis was performed in IBM SPSS Statistics version 27 using Dunnett’s post-hoc correction.

##### SNORD90 over-expression in mice

Male CD-1 (ICR) were housed in temperature-controlled (23 ± 1°C), constant humidity (55 ± 10%), 12-hour light/dark cycle and in specific-pathogen-free conditions. Animals had access to food and water *ad libidum*. Animals were housed in groups of four. All animal experiments were evaluated and approved by the local commission for the Care and Use of Laboratory Animals of the Government of Upper Bavaria, Germany.

#### Cloning

The Snord90 sequence was obtained from the Ensembl genome browser database (Ensemble ID ENSMUSG00000077756). The control sequence was generated by scrambling the Snord90 gene using siRNA wizard software (Invitrogen). Each fragment was designed to have KpnI and BamHI and a 10-nucleotide long overhang with the vector backbone at the 5’ and 3’ ends respectively. Each gene fragment was inserted into a KpnI with BamHI linearized pAAV-EF1a-eGFP-H1 backbone using Gibson Assembly (NEB) according to the manufacturer’s protocol.

This generated the following two vectors, pAAV-EF1a-eGFP-H1-Scramble and pAAV-EF1a-eGFP-H1-Snord90. All plasmids were checked for mutations by DNA sequencing.

SNORD90 sequence (mouse):

5’AAATAATGTTTTTAAGTGTCTAGTGATGAATTTCATAGGGCAGATTCTGAGGTGAAAATTTAGTTCATCATTGATTGTCCTATTATGAAATCTGAAGACACTTGAAAACTA 3’

Scrambled sequence (mouse): 5’GATATTATAACGATTGTTGTATAATCTATTAGAGTGTATCGTAGGCAATGGCTATATGTAAATGTAACGTATTGTACTCGTATGATTAATTACATAATACGAATACGCTAG 3’

#### Validation of constructs

Mouse neuroblastoma neuro2a (N2a) cells were maintained at 37°C with 5% CO_2_ in Minimum Essential Medium (MEM), 1x Glutamax, supplemented with 1x non-essential amino acids, 1mM sodium pyruvate, 100 U/ml penicillin, 100 μg/ml streptomycin and 10% fetal bovine serum (FBS, Gibco). Cells were detached with trypsin and transfected using ScreenfectA (ScreenFect GmbH) according to the manufacturer’s protocol. Cells were fixed with 4% PFA-PBS solution and embedded with Fluoromount-G mounting medium containing DAPI (SouthernBiotech). Cells were imaged using an Axioplan 2 fluorescent microscope (Zeiss).

#### Virus production

Human embryonic kidney cells (HEK293) were cultured in Dulbecco’s Modified Eagle Medium (DMEM) supplemented with 10% FBS, 100 U/ml penicillin and 100 μg/ml streptomycin (Invitrogen) in a 5% CO_2_ humidified incubator at 37°C. Cells were transfected with the gene transfer rAAV plasmid combined with the helper plasmids in an equal molar ratio of 1:1:1 using 1 mg/ml linear polyethylenimine hydrochloride (PEI). The rAAV (serotype 1/2) particles were harvested three days after transfection by lysing the cells with three consecutive freeze-and-thaw cycles using an ethanol on dry ice bath and 37°C water bath. Lysates were centrifuged (3000 rcf) followed by purification of the rAAV particles using a Heparin Agarose Type I chromatography column (Sigma). The eluted rAAV particles were PBS washed using a 100000 MWCO Amicon Ultra Filter (Millipore) and suspended in a final volume of 100μl. The number of viral genomic particles was determined using quantitative RT-PCR resulting in the following titers; AAV1/2-EF1a-eGFP-H1-Scramble 1,67x1011 genome particles (gp)/µl and AAV1/2-EF1a-eGFP-H1-Scramble 5x1010 gp/µl.

#### Stereotactic surgery

Eight-week-old mice were anesthetized with isoflurane and placed in the stereotactic apparatus (TSE Systems) on a 37°C heating pad. Pre-surgery, mice were given Novalgin (200 mg/kg body weight) and Metacam (sub-cutaneous 0.5 mg/kg body weight). During surgery, mice were continuously supplied with 2% v/v isoflurane in O_2_ through inhalation. Viruses were injected bilaterally using a 33-gauge injection needle with a 5 µl Hamilton syringe coupled to an automated microinjection pump (World Precision Instruments). Virus was delivered at a rate of 0.1 µl/min, to inject 0.25 µl for behavior experiments, and 0.5 µl for molecular experiments. The injection coordinates were determined using the Franklin and Paxinos mouse brain atlas, from bregma: ML +/-0.3 mm bilateral; AP +1.2 mm; DV -1.8 mm. After injection the needle was retracted 0.01 mm and kept at the site for 2.5 minutes, followed by slow withdrawal. In all experiments, each group consisted of two scramble control virus injected and two Snord90 overexpression virus injected animals. After surgery, the animals received Metacam for the 3 following days (intraperitoneal 0.5 mg/kg body weight). In all experiments, mice were tested or tissue was extracted 3-4 weeks after surgery. After completion of the experiments, mice were sacrificed by isoflurane overdose. For imaging of brain material, the brains were removed and fixed in 4% PFA-PBS followed by dehydration in 30% sucrose-PBS solution for at least 24 hours each. Brains were sectioned (50 μm) using a vibratome (HM 650 V, Thermo Scientific). Brain slices were imaged using the VS120-S6-W slide scanner microscope (Olympus). Injection sites were verified based on green fluorescent protein (GFP) expression. For RNA extraction, the brains were extracted and snap-frozen using methylbutane. Brains were sectioned in 200 μm thick slices in a cryostat and the injection site was collected using 0.8 mm thick puncher. The tissue was stored at -80°C until RNA extraction.

#### Real-time PCR (RT-PCR)

Total RNA was extracted from cells or tissue using the miRNeasy Mini Kit (Qiagen). cDNA was generated using the high-capacity cDNA RT kit with RNase inhibitor (Applied Biosystems) according to the supplied protocol. RT-PCR was performed according to the manufacturer’s instructions using the QuantiFast SYBR green PCR Kit (Qiagen). RT-PCR data was collected on the QuantStudio 7 Flex Real-Time PCR System (Applied Biosystems). Absolute expression differences were calculated using the standard curve method. See supplementary table 8 for a list of primer sequences used.

#### Behavioral tests

For all behavioral tests, animals were brought into the room 30 minutes prior to the start of the test to habituate the animals to the test room.

Open field (OF) test: The OF test was performed in a 50 x 50 cm light grey box, evenly illuminated with low light conditions (<10 lux). Mice were placed in the open field facing one of the walls and recorded for 6 minutes. Animals were tracked using ANY-maze software (Stoelting). Total distance traveled was used as measure of animal locomotion.

Elevated plus maze (EPM): EPM apparatus was made of light grey material, and consists of four intersecting arms elevated approximately 30 cm above the floor. The two opposing open (27.5 x 5 cm) and closed arms (27.5 x 5 x 20 cm) are connected with a central zone (5 x 5 cm). The animals were placed in the center of the EPM, facing one of the open arms and recorded for 6 minutes. The closed arms were illuminated with <10 lux, while the open arms were illuminated with 25-30 lux. Recordings were tracked and analyzed with ANYmaze software. Total time spent in the open or closed arms was used as an anxiety measure.

Splash test (Spl): Animals were placed in a novel cage containing fresh bedding material and were allowed to explore for 5 minutes. Each animal received two sprays of room temperature 10% sucrose water at the rear of their body. Animals were recorded for 6 minutes under low-light conditions (<10 lux). The total groom time within the 3-6 minute timeframe was manually scored using Solomon Coder software.

Tail suspension test (TST): In the TST, animals were taped by their tales on a metal rod, approximately 30 cm above the ground, and illuminated with 30-35 lux. Animals were recorded for 6 minutes and struggle time was quantified using ANY-maze software. The total time an animal struggled within the 3-6 minute timeframe was used as a measure of a depression-like emotional state.

Z-scoring was used to integrate multiple behavioral tests as previously described (*45*). First, the z-score of each individual behavioral parameter was calculated.

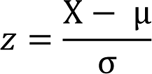

The z-scores of the EPM open arm, EPM closed arm, Spl and TST were combined to calculate an integrated emotionality score.

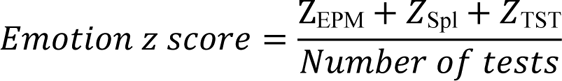

The final integrated score is associated with anxiety and depressive-like behaviors with a higher score indicating a higher emotional state, while a lower score indicating a lower emotional state.

##### Target Prediction

The entire sequence of SNORD90 was used to blast against the human genome allowing G-T wobble base pairing and one mismatch base pairing (supplemental table 4). Additionally, the C/D box snoRNA target prediction algorithm PLEXY, using default parameters, was used for target prediction of SNORD90 (*17*). SNORD90 mature sequence was used as the input snoRNA and target sequence input was the entire human transcriptome using both mRNA and pre-mRNA sequences downloaded from the Ensembl genome browser database (supplementary table 5). Prediction was conducted using whole transcriptomics as well as a targeted analysis for NRG3.

##### SNORD90 in-vitro knock-down

SNORD90 was knocked down using antisense oligonucleotides (ASO) containing 2′-O-methylations and phosphorothioate-modified nucleotides (supplementary table 9). Four ASOs and one scrambled control ASO were screened to identify which achieved the best knock-down of SNORD90 (supplementary figure 4). NPCs were plated in 24-well plates until ∼90% confluent before transfection with ASO (50nM final concentration) using Lipofectamine 2000 (ThermoFisher) following manufacturer’s protocol. NPCs were incubated for 48 hours before harvesting for RNA extraction. RNA extraction, cDNA synthesis, and RT-qPCR were as described above. One-way ANOVA was used as described above

##### SNORD90 in-vitro over-expression

Constructs and cloning were as described above with the following sequences: SNORD90 sequence:

5’TTATAAGTTTTCTAAGTGTCTAATGATGAATTTCATAGGGCAGATTCTGAGGTGAAAATTTAATTCATCACTGATACTCCTACTGTGGAATCTGAAGACACTTGAAAACGT 3’

Seed Scrambled sequence: 5’TTATAAGTTTTCTAAGTGTCTAATGATGAATTTCATAGGGCAGATTCTGAAATAATACTTCGCTTAAGATATTCGACTCCTACTGTGGAATCTGAAGACACTTGAAAACGT 3’

Scrambled sequence: 5’GATTCATAATGAGTTGATTTAATCACATGCTGGCTCTCATTCGACCAGAATTTCTCTAGTTGATAAAAGTAAACCAATGAATTAGTATGTTTGATCAAGATGTATGACTCG 3’

NPCs were plated in 24-well plates until ∼90% confluent before being transfected with each vector using Lipofectamine 2000 (ThermoFisher) following manufacturer’s protocol. NPCs were incubated for 48 hours before harvesting for RNA extraction. RNA extraction, cDNA synthesis, and RT-qPCR were as described above. One-way ANOVA was used as described above.

#### Target Blockers

Modified RNA oligos containing 2′-O-methylations and phosphorothioate-modified nucleotides (target blockers) were designed against three regions on the NRG3 transcript where SNORD90 is predicted to bind (Figure 3A). Target blockers were co-transfected (30nM final concentration) with SNORD90 OE vectors using lipofectamine 2000 following manufacturer recommendations for “plasmid DNA and siRNA” co-transfection. For groups where target blockers were pooled together, equal amounts for each target blocker were used totaling 30nM final concentration.

NPC culture conditions, RNA extraction, cDNA synthesis, and RT-qPCR were as described above. One-way ANOVA was used as described above.

## Western blot

Cell pellets from cell culture were lysed in lysis buffer [150 mM NaCl, 50 mM HEPES (pH 7), 50 mM EDTA, and 0.1% NP-40] and quantified using PierceTM BCA Protein Assay Kit (Thermo Fisher). Equal amounts of proteins were electrophoresed on 4-20% Mini-PROTEAN TGX Stain-Free Gels (Bio Rad). Proteins were then transferred onto nitrocellulose membranes using the Trans-Blot Turbo Transfer System (Bio Rad). The membranes were blocked with 5% Bovine Serum Albumin (BSA) in Phosphate-Buffered Saline (0.05% Tween 20) (PBS-T) at room temperature for 1-2 hrs and then incubated with primary antibody (NRG3 (ab109256) at 1:500 and GAPDH (3683S) at 1:1000) in 1% BSA in PBS-T overnight at 4 oC. They were then incubated with either biotin-conjugated anti-rabbit antibody (BA-1000) or with Horseradish Peroxidase (HRP)-conjugated anti-rabbit antibody diluted 1:5000 in 1% BSA in PBS-T for 1 hr at room temperature. The membranes that were incubated with biotin-conjugated anti-rabbit antibody were washed with PBS-T and incubated with streptavidin-conjugated HRP (016-030-084) at 1:5000 in 1% NFDM in PBS-T for 1 hr at room temperature. After washing, immunoreactivity was detected using enhanced chemiluminescence solutions (ECL) and the Biorad ChemiDocTM MP imaging system. Western blots were performed in 3 biological replicates.

### RNA immunoprecipitation

Overexpression of SNORD90 and scramble were performed as described above. RNA immunoprecipitation (RIP) was performed using the Magna RIP RNA-Binding Protein Immunoprecipitation kit (Millipore Sigma) following the manufacturer’s protocol with slight modifications. NPCs were collected and lysed in complete RIP lysis buffer. 10 uL of cell extract was stored at -80°C and used as input. NPC extracts were incubated in RIP buffer containing magnetic beads conjugated to anti-fibrillarin antibody (ab226178; abcam), anti-RBM15B antibody (22249-1-AP; proteintech), or IgG control (provided by the kit) overnight in 4°C. Bead complexes were washed 6 times with RIP buffer and eluted directly in TRIzol. RNA from all conditions were purified in parallel using the RNA microPrep kit (Zymol) and eluted into 15 μL H2O. The entire eluate was transcribed to cDNA using M-MLV Reverse Transcriptase (200 U/µL) (ThermoFisher) with random hexamers. RT-qPCR was as described above.

### m6A-RIP

m6A-RIP protocol was as described by Engel et al., with minor modifications. 3 μg total RNA was mixed with 3 fmol spike-in and equally split into 3 conditions: m6A-RIP, IgG control and input. Input samples were frozen and kept at -80C during the m6A-RIP protocol. m6A-RIP and IgG control samples were incubated with 1 μg anti-m6A antibody (rabbit polyclonal 202 003, Synaptic Systems) or 1 μg normal rabbit IgG (NEB) in immunoprecipitation (IP) buffer (10 mM Tris-HCl [pH 7.5], 150 mM NaCl, 0.1% IGEPAL CA-630 in nuclease-free H2O, 0.5 mL total volume) with 1 μL RNasin Plus (Promega) rotating head over tail at 4°C for 2 hrs, followed by incubation with 2x washed 25 μL Dynabeads M-280 (Sheep anti-Rabbit IgG Thermo Fisher Scientific) rotating head over tail at 4°C for 2 hrs. Bead-bound antibody-RNA complexes were recovered on a magnetic stand and washed in the following order: twice with IP buffer, twice with high-salt buffer (10 mM Tris-HCl [pH 7.5], 500 mM NaCl, 0.1% IGEPAL CA-630 in nuclease-free H2O), and twice with IP buffer. RNA was eluted directly into TRIzol and input RNA was also taken up in TRIzol. RNA from all conditions was purified in parallel using the RNA microPrep (Zymol) and eluted into 15 μL H2O. The entire eluate was transcribed to cDNA using M-MLV Reverse Transcriptase (200 U/µL) (ThermoFisher) with random hexamers. RT-qPCR was as described above. One-way ANOVA was used as described above

#### Spike-in

A spike-in Oligo was used as a normalizer for quantitative RT-PCR. The spike-in oligo was 100nt in length with 3 internal m6A/m sites within GGAC motif flanked by the most frequent nucleotides 5′ U/A, 3′ A/U, not complementary to hsa or mmu RefSeq mRNA or genome, secondary structure exposing m6A sites (as described by Engel et al., 2018). The sequence is: GCAGAACCUAGUAGCGUGUGGmACACGAACAGGUAUCAAUAUGCGGGUAUGGmA CUAAAGCAACGUGCGAGAUUACGCUGAGGmACUACAAUCUCAGUUACCA (synthesized by Horizon Discoveries).

### Dicer-Substrate Short Interfering RNA (dsiRNA) gene silencing

DsiRNAs KD was assessed via RT-qPCR in the final experimental samples (supplementary figure 8). In each silencing condition, the siRNA sequences were specific to the m6a reader of interest, as shown by the quantitative RT-PCR data (supplementary figure 8). Cells were seeded in 24-well plates and grown to ∼70% confluency. 50nM siRNA was transfected using lipofectamine 2000 according to the manufacturer’s instructions. A second transfection was performed 48hrs after the first transfection. Following the second round of dsiRNA transfection, SNORD90 OE vectors were transfected 48 hours later. NPCs were then collected 48 hours after the third transfection. DsiRNA sequences and oligos were provided by IDT (supplementary table 10). RNA extraction, cDNA synthesis, and RT-qPCR were as described above. One-way ANOVA was used as described above

### Electrophysiological Recording

#### Brain slices preparation

Mice were stereotactically injected with SNORD90-OE or SNORD90-Scramble viral vector as described above.

3 weeks after the injection, mice received an overdose injection of pentobarbital (100 mg/kg i.p.) and were perfused with carbogenated (95% O_2_, 5% CO_2_) ice-cold slicing solution containing (in mM): 2.5 KCl, 11 glucose, 234 sucrose, 26 NaHCO3, 1.25 NaH2PO4, 10 MgSO4, 2 CaCl2; pH 7.4, 340 mOsm. After decapitation, 300 μm coronal slices containing the ACC were prepared in carbogenated ice-cold slicing solution using a vibratome (Leica VT 1200S) and allowed to recover for 20 min at 33°C in carbogenated high osmolarity artificial cerebrospinal fluid (high-Osm aCSF) containing (in mM): 3.2 KCl, 11.8 glucose, 132 NaCl, 27.9 NaHCO3, 1.34 NaH2PO4, 1.07 MgCl2, 2.14 CaCl2; pH 7.4, 320 mOsm) followed by 40 min incubation at 33°C in carbogenated aCSF containing (in mM): 3 KCl, 11 glucose, 123 NaCl, 26 NaHCO3, 1.25 NaH2PO4, 1 MgCl2, 2 CaCl2; pH 7.4, 300 mOsm. Subsequently, slices were kept at room temperature (RT) in carbogenated aCSF until use.

#### Patch-clamp recordings

Slices were then transferred in the recording chamber and superfused (4-5 mL/min) with carbogenated aCSF and recordings performed at RT. Pyramidal neurons of the ACC were visualized with infrared differential interference contrast (DIC) microscopy (BX51W1, Olympus) and an Andor Neo sCMOS camera (Oxford Instruments, Abingdon, UK). Somatic whole-cell voltage-clamp recordings from ACC pyramidal neurons (> 1 GΩ seal resistance, -70 mV holding potential) were performed using a Multiclamp 700B amplifier (Molecular Devices, San Jose, CA, USA). Data were acquired using pClamp 10.7 on a personal computer connected to the amplifier via a Digidata-1440 interface (sampling rate: 20 kHz; low-pass filter: 8 kHz). Borosilicate glass pipettes (BF100-58-10, Sutter Instrument, Novato, CA, USA) with resistances 4-6 MΩ were pulled using a laser micropipette puller (P-2000, Sutter Instrument). Data obtained with a series resistance > 20 MΩ or fluctuation more than 20% of the initial values were discarded.

For sEPSCs recording, pyramidal neurons were clamped at -70 mV in the presence of BIM (20 uM) with the pipette solution containing (in mM): 125 Cs-methanesulfonate, 8 NaCl, 10 HEPES, 0.5 EGTA, 4 Mg-ATP, 0.3 Na-GTP, 20 Phosphocreatine and 5 QX-314 (pH 7.2 with CsOH, 285-290 mOsm).

Analysis was performed using ClampFit 10.7 and Easy Electrophysiology V2.2 (https://www.easyelectrophysiology.com), and statistical significance assessed with GraphPad Prism 7.

## Data Availability

The raw sequencing data generated in this study are available upon request on GEO using accession number (TBD). qPCR data are available upon request.

## ACKNOWLEDGMENTS

GT holds a Canada Research Chair (Tier 1) and is supported by grants from the Canadian Institute of Health Research (CIHR) (FDN148374, EGM141899, ENP161427), and by the Fonds de recherche du Québec -Santé (FRQS) through the Quebec Network on Suicide, Mood Disorders, and Related Disorders. CAN-BIND is an Integrated Discovery Program carried out in partnership with, and financial support from, the Ontario Brain Institute, an independent nonprofit corporation, funded partially by the Ontario government. The opinions, results, and conclusions are those of the authors and no endorsement by the Ontario Brain Institute is intended or should be inferred. Additional funding is provided by the Canadian Institutes of Health Research (CIHR). All study medications were independently purchased at wholesale market values. AC is the incumbent of the Vera and John Schwartz Family Professorial Chair in Neurobiology at the Weizmann Institute and the head of the Max Planck Society–Weizmann Institute of Science Laboratory for Experimental Neuropsychiatry and Behavioral Neurogenetics. This work is supported by the German Ministry of Science and Education (IMADAPT, FKZ: 01KU1901); the Ruhman Family Laboratory for Research in the Neurobiology of Stress (AC); research support from Bruno and Simone Licht; the Perlman Family Foundation, founded by Louis L. and Anita M. Perlman (AC); the Adelis Foundation (AC); and Sonia T. Marschak (AC). JPL holds postdoctoral fellowships from the European Molecular Biology Organization (EMBO-ALTF 650-2016), Alexander von Humboldt Foundation, and the Canadian Biomarker Integration Network in Depression (CAN-BIND). J.D. is the incumbent of the Achar Research Fellow Chair in Electrophysiology.

## AUTHOR CONTRIBUTIONS

RL and GT conceptualized the project with input from AC, JPL AK, and LMF. JPL performed small RNA sequencing from human clinical samples. PB, FF, BNF, RWL, RM, DJM, SVP, CS, RU, JAF, and SHK participated in the design the sample accusation of the human clinical trials. RL and LMF performed small RNA sequencing on human post-mortem brain samples. CN and NM contributed to the collection of the human-postmortem samples and molecular analysis. ZA provided bioinformatic support for analysis of sequencing data. AK performed mouse behavioral experiments. ECI and CB provided mouse ACC samples. JD performed electrophysiology experiments; YJB performed surgeries with input from AK and JPL. TPW provided input on electrophysiology experiments. PI performed western blots. RL performed cell culture experiments with input from JY. RL and HH performed m6A experiments. RL and GT wrote the manuscript with input from AK, JPL, JD, JBF, TPW, ECI, CB, PB, FF, BNF, RWL, RM, DJM, SVP, CS, RU, CN, NM, JAF, SHK, and AC.

## Supplementary Materials

**This PDF file includes:** Figs. S1 to S10

**Supplementary Figure 1:**
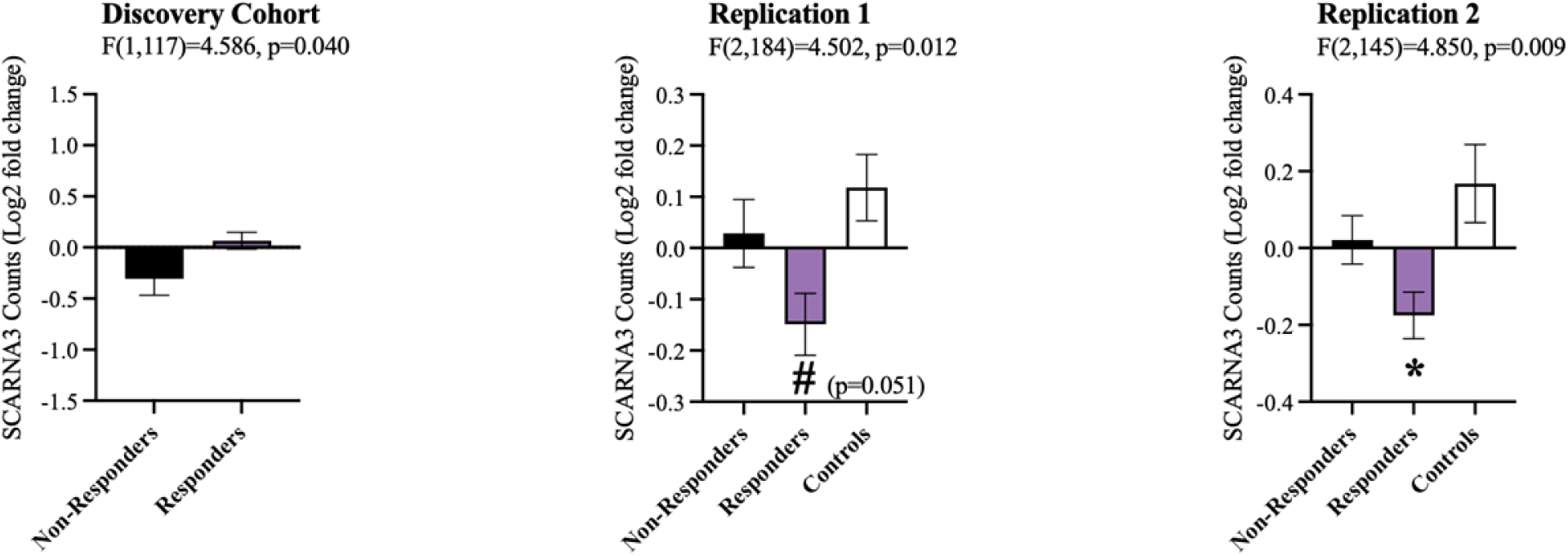
SCARNA3 expression in human clinical cohorts (related to figure 1) Log2 fold-change of the expression of SCARNA3 before and after antidepressant treatment for all three clinical cohorts. SCARNA3 displayed inconsistent expression after eight weeks of antidepressant treatment.

**Supplementary Figure 2:**
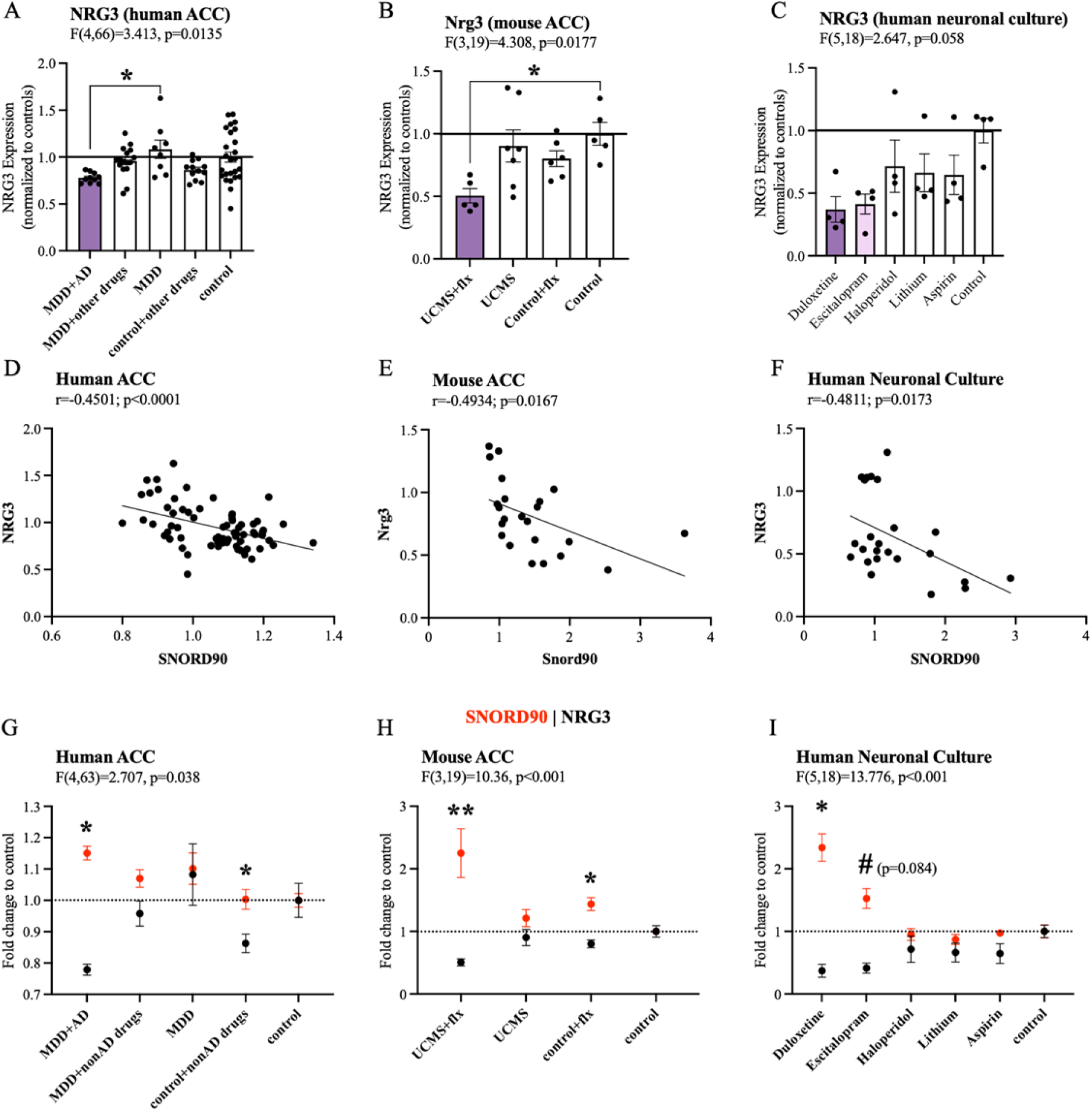
NRG3 expression negatively correlates with SNORD90 in the context of antidepressant (related to figure 1) **A** Nrg3 expression in the ACC of mice that underwent unpredictable chronic mild stress (UCMS) and antidepressant administration. **B** NRG3 expression in human post-mortem ACC. Samples were separated based on presence or absence of antidepressant drug treatment. **C** NRG3 expression in human neuronal cultures exposed to various psychotropic drugs. **D-F** Correlation between SNORD90 and NRG3 expression in each of the respective experiments listed in A-C. **G-I** SNORD90 (red) and NRG3 (black) fold change in relation to control conditions. SNORD90 and NRG3 displayed the most opposing direction of expression in groups exposed to antidepressants. **A-C** Statistical analysis using one-way ANOVA with Bonferroni post-hoc. **D-F** Pearson correlation. **G-I** Statistical analysis using two-way mixed ANOVA with Bonferroni post-hoc. All bar plots represent the mean with individual data points as dots. Error bars represent S.E.M. (*p<0.05, **p<0.01).

**Supplementary Figure 3:**
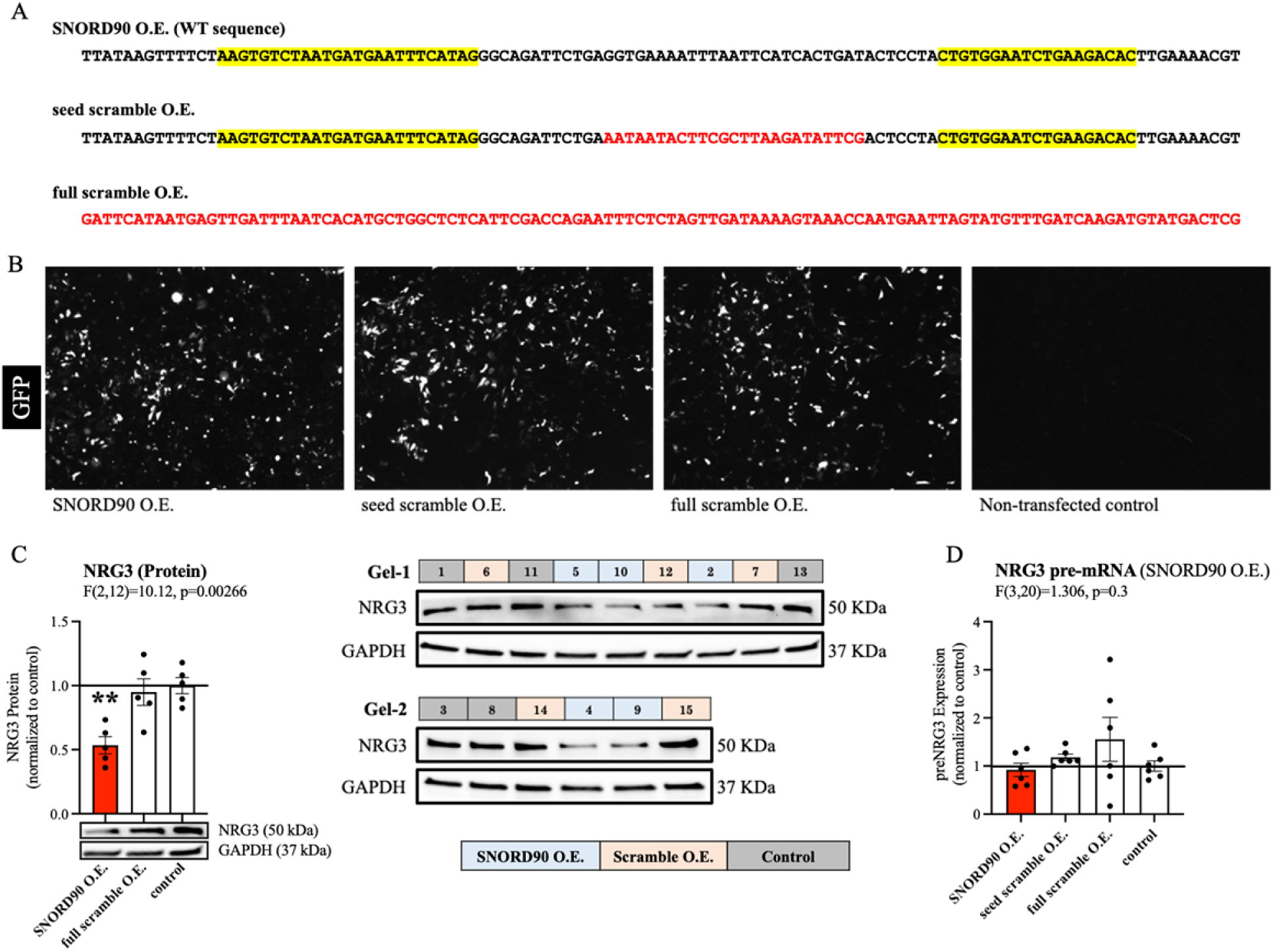
SNORD90 over-expression in human NPCs (related to figure 3) **A** Sequences of each over-expressed transcript. Red indicates scrambled sequence and black indicates wild-type sequence. Yellow highlight indicates where forward and reverse primers bind for qPCR quantification. Over-expression of SNORD90 WT transcript and seed scramble will be detected by our primers however full scramble will not. **B** Image of NPC 48hrs post-transfection. White is the expression of GFP to confirm successful transfection. **C** Quantification of NRG3 protein levels following SNORD90 over-expression. NRG3 protein levels were significantly reduced after SNORD90 over-expression (left). Western blots where samples were equally distributed between two separate gels and randomly assigned a well (right). **D** Expression of NRG3 pre-mRNA following SNORD90 over-expression. **C-D** Statistical analysis using one-way ANOVA with Bonferroni post-hoc. All bar plots represent the mean with individual data points as dots. Error bars represent S.E.M. (*p<0.05, **p<0.01). **Supplementary Figure 3C-source data** Source data for western blots for supplementary figure 3C including original unedited files of each western blot.

**Supplementary Figure 4:**
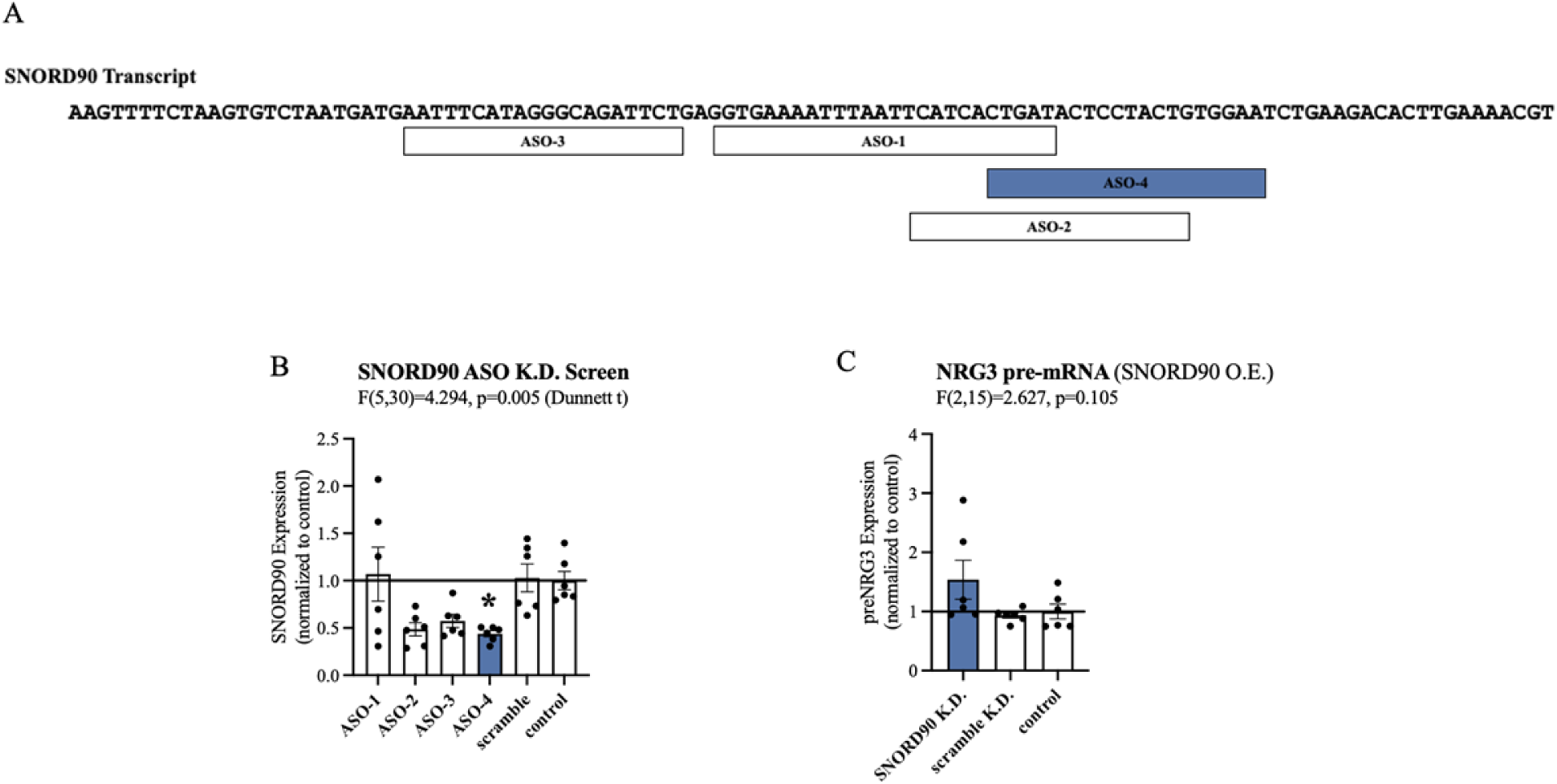
SNORD90 knock-down in human NPCs (related to figure 3) **A** Schematic diagram indicating where each ASO is designed to bind onto SNORD90. **B** Screen of all four ASOs to determine which ASO produced the best knock-down of SNORD90. **C** Expression of NRG3 pre-mRNA after SNORD90 knock-down. **B-C** Statistical analysis using one-way ANOVA with Bonferroni post-hoc (unless otherwise indicated on the graph). All bar plots represent the mean with individual data points as dots. Error bars represent S.E.M. (*p<0.05).

**Supplementary Figure 5:**
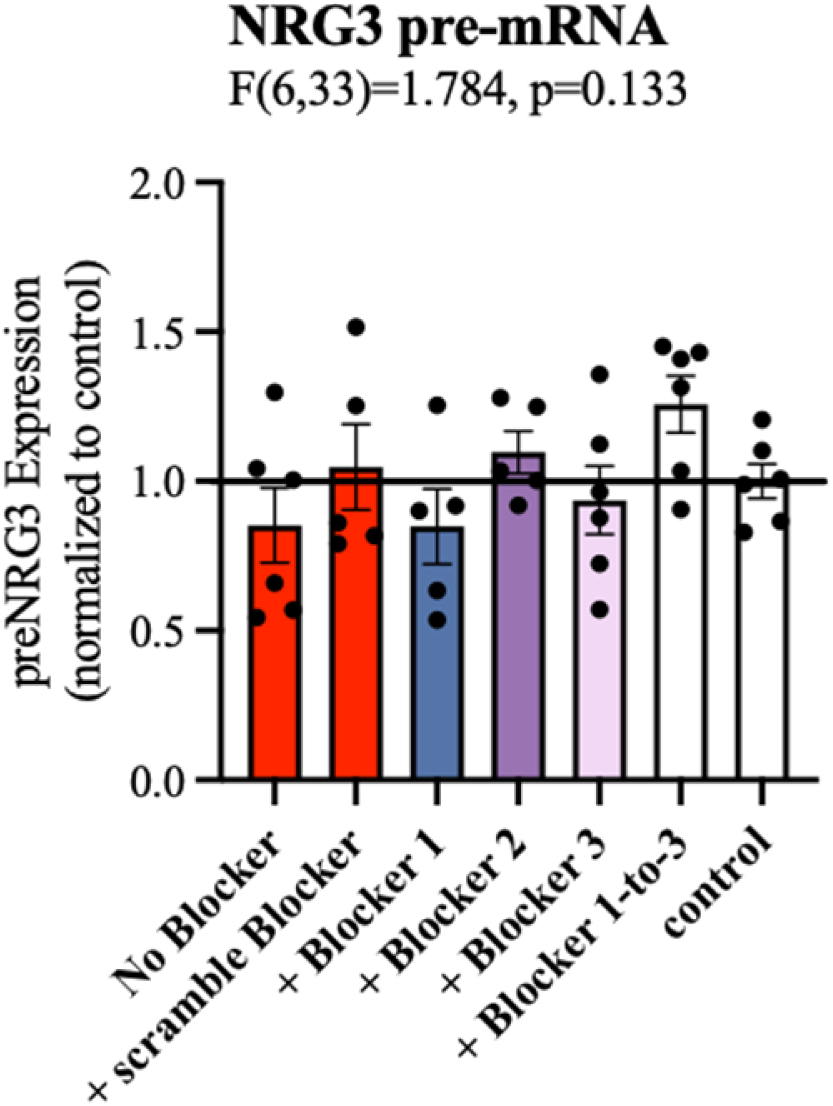
SNORD90 over-expression and NRG3 target blockers (related to figure 3) Expression of NRG3 pre-mRNA after SNORD90 over-expression followed by NRG3 target blockers. Statistical analysis using one-way ANOVA with Bonferroni post-hoc (unless otherwise indicated on the graph). All bar plots represent the mean with individual data points as dots. Error bars represent S.E.M.

**Supplementary Figure 6:**
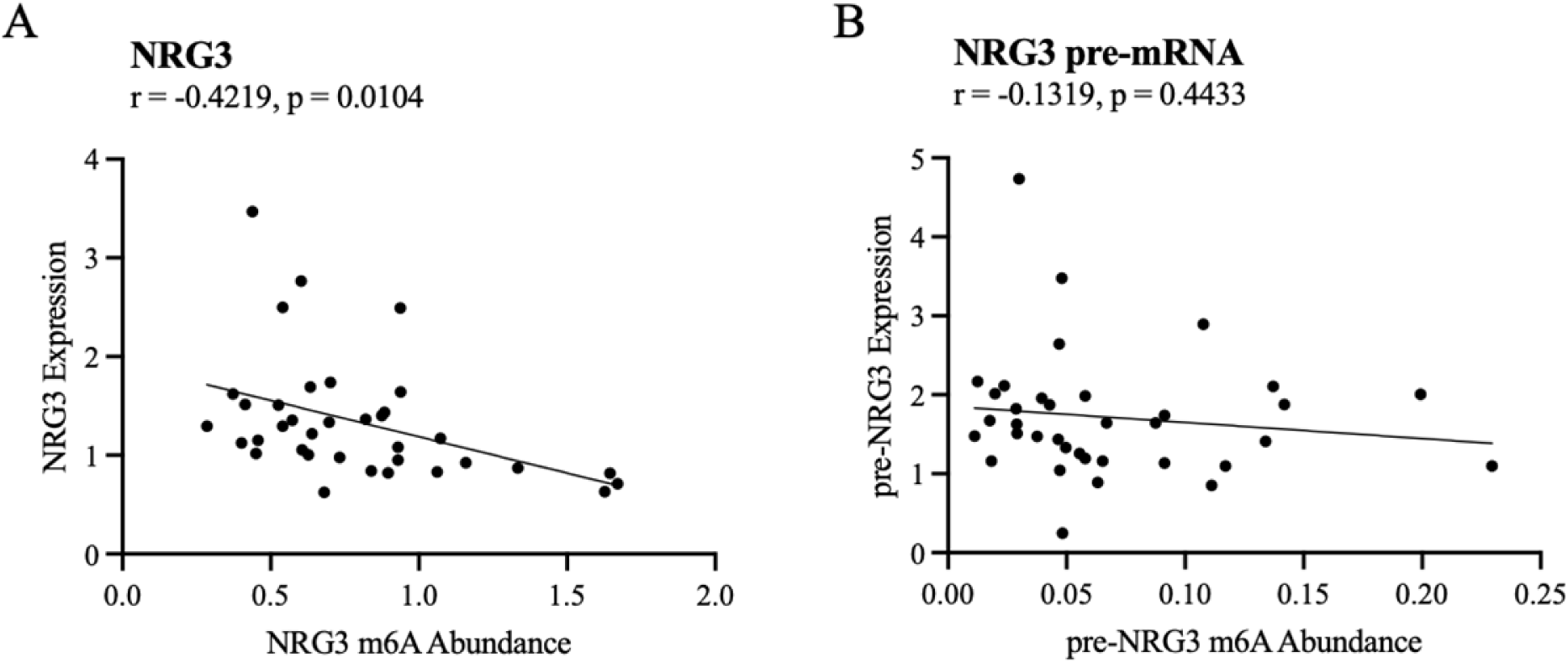
m6A abundance on NRG3 is related to expression (related to figure 4) **A** Correlation of total m6A abundance on NRG3 and NRG3 expression. **B** Correlation of total m6A abundance on NRG3 pre-mRNA and NRG3 pre-mRNA expression. **A-B** person correlation

**Supplementary Figure 7:**
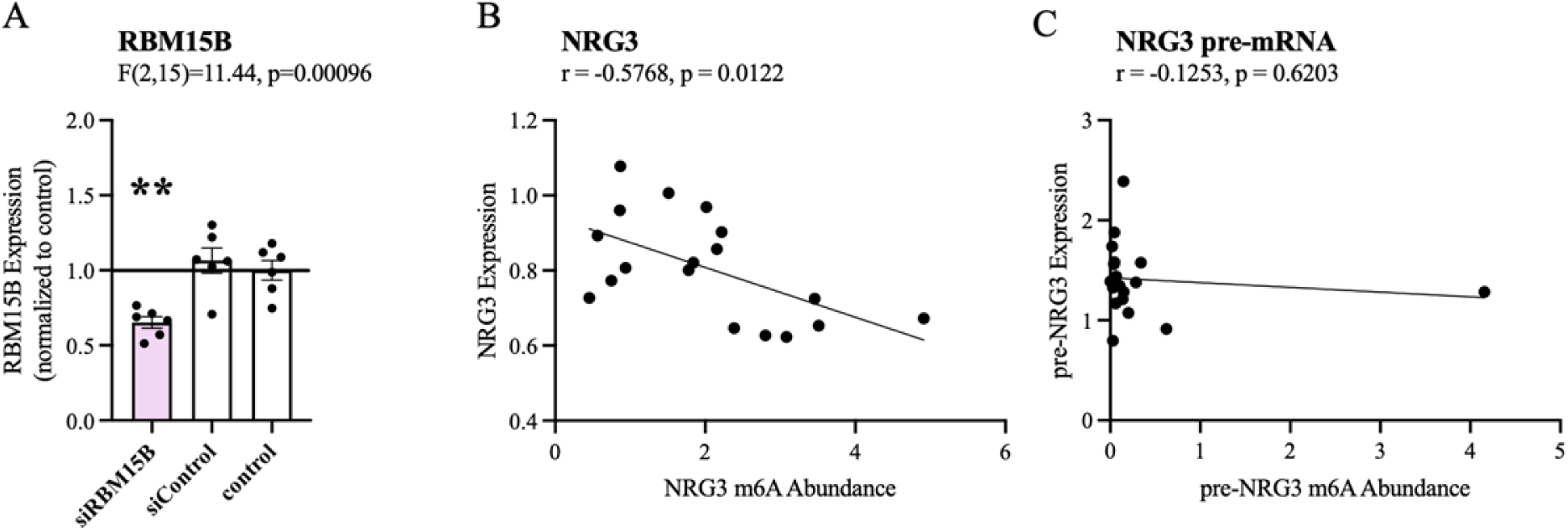
RBM15B knock-down (related to figure 4) **A** qPCR validation of RBM15B knock-down using dsiRNAs. **B-C** Correlation of total m6A abundance on NRG3 or NRG3 pre-mRNA and expression. **B-C** Person correlation. **A** Statistical analysis using one-way ANOVA with Bonferroni post-hoc. All bar plots represent the mean with individual data points as dots. Error bars represent S.E.M. (**p<0.01).

**Supplementary Figure 8:**
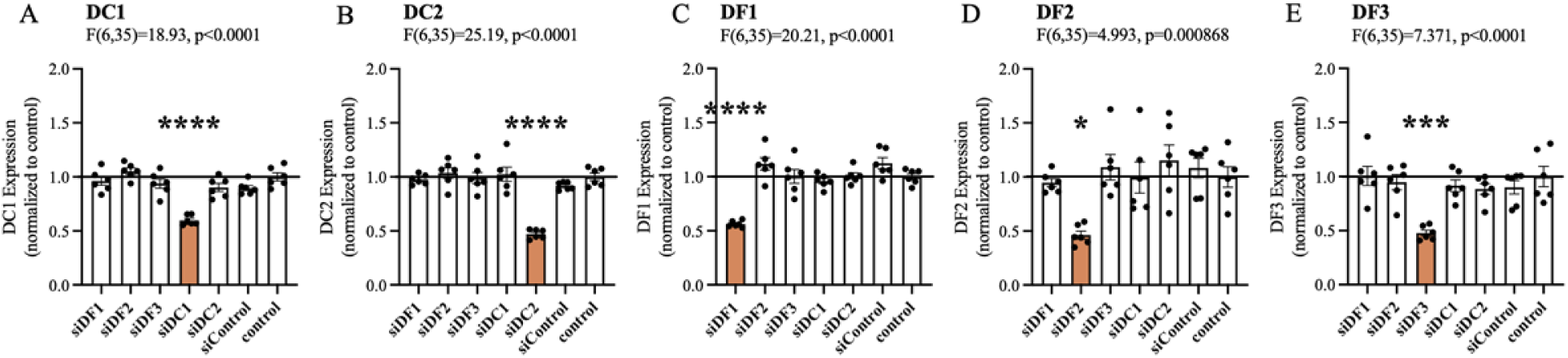
m6A-reader knock-down (related to figure 4) **A-E** qPCR validation of each respective m6A-reader knock-down using dsiRNAs. **A-E** Statistical analysis using one-way ANOVA with Bonferroni post-hoc. All bar plots represent the mean with individual data points as dots. Error bars represent S.E.M. (* p<0.05, **p<0.01, ***p<0.001, ****p<0.0001).

**Supplementary Figure 9:**
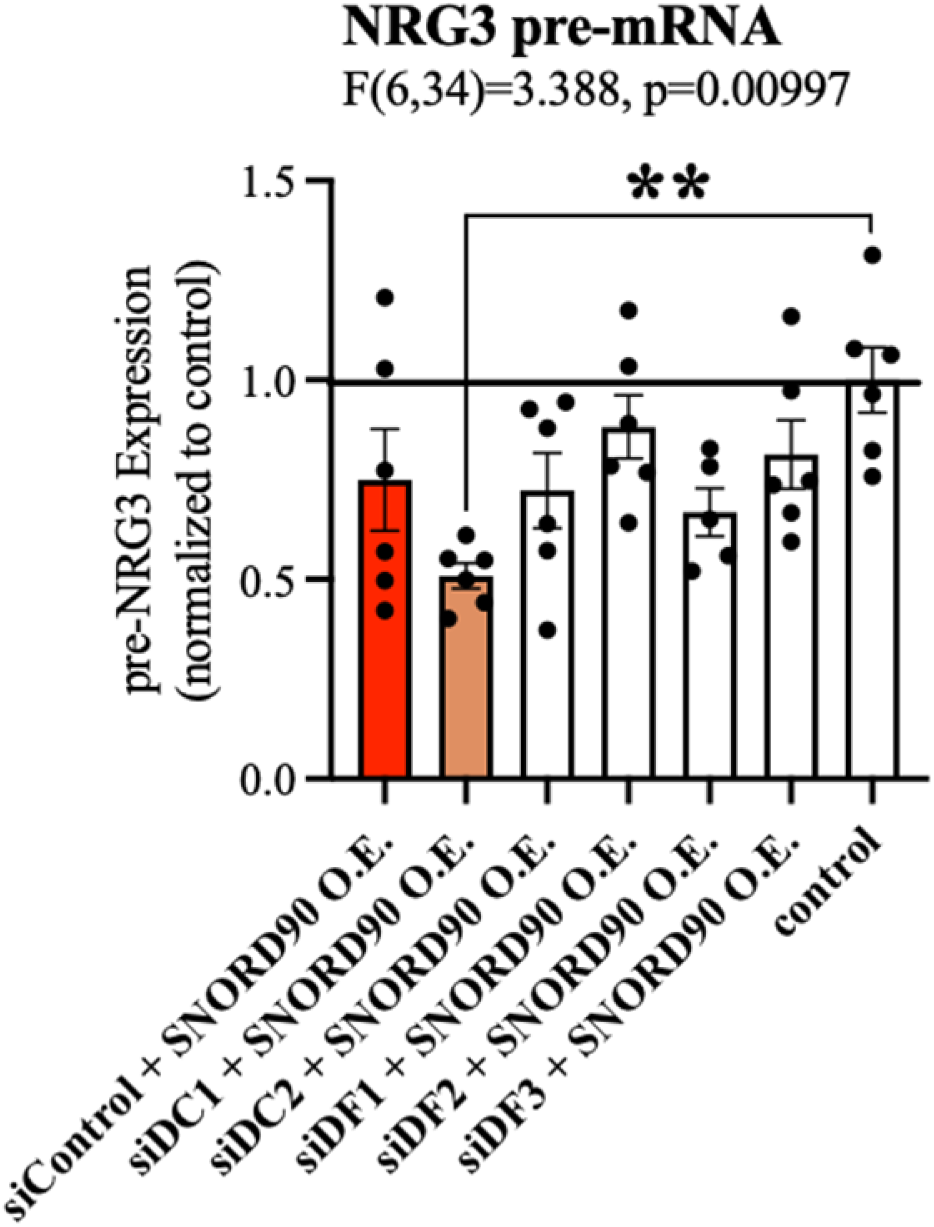
Expression of NRG3 pre-mRNA after m6A-reader knock-down (related to figure 4) Expression of NRG3 pre-mRNA following m6A-reader knock-down and SNORD90 over-expression. Statistical analysis using one-way ANOVA with Bonferroni post-hoc. All bar plots represent the mean with individual data points as dots. Error bars represent S.E.M. (**p<0.01).

**Supplementary Figure 10:**
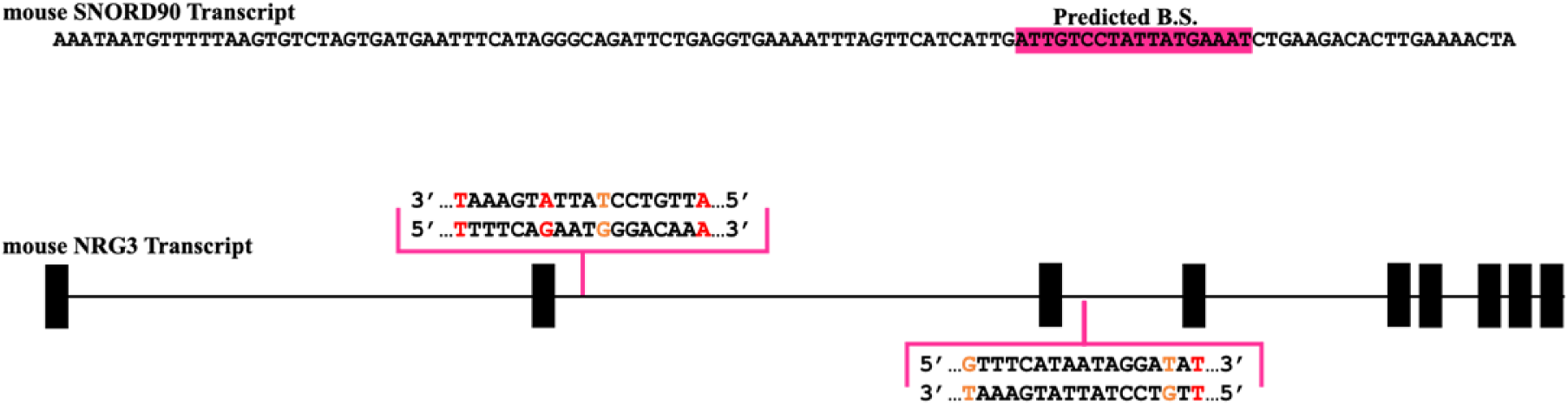
Snord90-nrg3 interaction is conserved in mice (related to figure 5) Schematic diagram showing predicted interaction sites between Snord90 and Nrg3 in mice

**Other Supplementary Material for this manuscript includes the following:** Table S1 to S10 (.xlsx)

Supplementary Table 1 to 10

**Supp. Table 1 (separate file).** Summary statistics for small RNA sequencing analysis in human clinical discovery cohort

**Supp. Table 2 (separate file).** Summary statistics for small RNA sequencing analysis in human clinical replication 1 cohort

**Supp. Table 3 (separate file).** Summary statistics for small RNA sequencing analysis in human clinical replication 2 cohort

**Supp. Table 4 (separate file).** SNORD90 BLAST alignment results

**Supp. Table 5 (separate file).** SNORD90 PLEXY target prediction results (human)

**Supp. Table 6 (separate file).** SNORD90 RNA binding protein motif prediction results

**Supp. Table 7 (separate file).** SNORD90 PLEXY target prediction results (mouse)

**Supp. Table 8 (separate file).** qPCR primer sequences

**Supp. Table 9 (separate file).** ASO sequences

**Supp. Table 10 (separate file).** dsiRNA sequences

